# Antimicrobial selection for resistance in four major pathogens in the US Veterans Affairs Healthcare System, 2007-2021

**DOI:** 10.1101/2025.03.12.25323875

**Authors:** Thi Mui Pham, Yue Zhang, McKenna Nevers, Haojia Li, Karim Khader, Yonatan H. Grad, Matthew Samore, Marc Lipsitch

## Abstract

**Background:** Systematic evidence on antimicrobial selection for antimicrobial resistance (AMR) is scarce. We estimated the effect of prescribing key antibiotic classes on AMR across U.S. Veterans Affairs Medical Centres (VAMC).

**Methods:** We analysed clinical isolates of *Staphylococcus aureus, Escherichia coli, Klebsiella pneumoniae*, and *Pseudomonas aeruginosa* from 138 VAMC from Feb 1, 2007 to Dec 31, 2021. Antimicrobial prescribing was measured as inpatient days of therapy per 1000 patient-days; multidrug resistance as number of resistant phenotypes per 1,000 admissions. Temporal trends were modelled using generalized estimating equations and average annual percentage changes (AAPC). Multilevel multinomial logistic regression related facility-level antibiotic prescribing (days of therapy per 100 patient-days in the last 14d) to the relative odds of resistant phenotypes.

**Findings:** Hospital-onset infection incidence declined for all pathogens, except third-generation cephalosporin (3GC)-resistant *E coli*. Antimicrobial prescribing remained stable or decreased, except 3GC prescribing, which increased from 2007 until 2019 (AAPC=2·4%, 95% CI 1·3%–3·5%, p-value<0.0001). Fluoroquinolone (FQL) use was associated with resistance across all pathogens. In *S aureus*, each day of FQL treatment was linked to a 4·6% (95CI: 1·5, 7·7, p-value=0.0127) increase in the relative odds of isolating FQL-resistant, macrolide-susceptible, methicillin-resistant *S aureus*. Anti-staphylococcal beta-lactams were not linked to MRSA. Each day of 3GC treatment increased the odds of isolating 3GC- and beta-lactam/beta-lactamase-resistant *E coli* by 5·2% (95%CI: 1·3%, 9·4%, p-value=0.0079) and *K pneumoniae* by 3·0% (95% CI: −0·1%-6·2%, p-value=0.0600). Each day of carbapenem treatment increased the odds of carbapenem-resistant, FQL- and BL/BLI-susceptible *P aeruginosa* by 15·7% (95%CI: 9·4%, 22·4%, p-value<0.0001).

**Interpretation:** Higher facility-level antimicrobial use increased the odds of corresponding resistant phenotypes, with important exceptions. FQLs selected for resistance across multiple pathogens. Increased 3GC prescribing likely offset reductions in FQLs and was associated with co-resistance in *E coli*. These findings underscore the need for comprehensive stewardship that coordinates strategies across antimicrobials.

## Introduction

Antimicrobial use has long been recognized as a major driver of antimicrobial resistance (AMR).^1^ However, our understanding of the complex relationship between antimicrobial use and resistance remains incomplete, partly because of methodological challenges in assessing this association.^2^ Many studies have demonstrated ecological correlations for single pathogen-single antimicrobial combinations at regional, national, or international scales.^3–10^ The evidence linking use to resistance is often contradictory, with some studies showing significant correlations for certain pathogens, while others reveal no clear association, particularly for commensal bacteria.^3–6^ A landmark study of 26 European countries used data from 1997 to 2002 and correlated the use of β-lactams and fluoroquinolones with resistance in *Streptococcus pneumoniae, S. pyogenes*, and *Escherichia coli* isolates.^7^ They showed that nations with the highest outpatient antimicrobial consumption had the greatest prevalence of resistant bacteria, whereas low-consumption countries had much lower resistance rates. In the United States, cross-sectional analyses have similarly examined multiple pathogen-antimicrobial pairs.^3,10^ Olesen and colleagues described the landscape of correlations for 72 pathogen-antimicrobial combinations across U.S. states using outpatient antimicrobial prescription data covering 2011-2014.^3^ The most positive significant correlations involved macrolides, fluoroquinolones, and cephalosporins. Another analysis of five pathogen-antimicrobial combinations in eight U.S. hospitals showed that the use of anti-staphylococcal agents and percentage of methicillin-resistant *Staphylococcus* aureus (MRSA) were not correlated.^10^

Few studies have investigated the potential impact of co-selection,^11^ where use of one antimicrobial agent promotes the selection and persistence of resistance not only to that agent but also to others, on the use-resistance relationship.^12–14^ In multivariate models for *S pneumoniae*, macrolide consumption explained as much variation in penicillin resistance as penicillin use, implying that macrolide exposure co-selected penicillin-resistant strains.^15^ Pouwels and colleagues found that increased prescribing of amoxicillin in the community was associated not only with more amoxicillin-resistant *E coli* in urine isolates, but also with higher ciprofloxacin and trimethoprim resistance in those *E coli*.^13^ These studies typically aggregate data at a high level (e.g., states, nations) and examine correlations between total antimicrobial consumption and overall resistance prevalence. Such analyses provide a broad view of the association but can mask within-group variability, may suffer from ecological fallacies, and are often subject to reverse confounding by not capturing the precise time lags between antimicrobial exposure and the development of resistance.^2^

Several other studies investigated the relationship between antimicrobial use and resistance on an individual-level.^16^ In a case-control study, Weber and colleagues found that exposure to fluoroquinolones, specifically levofloxacin and ciprofloxacin, was a significant risk factor for the subsequent isolation of MRSA in hospitalized patients, but not for methicillin-susceptible *S aureus* (MSSA), highlighting the role of fluoroquinolones in co-selecting MRSA.^17^ However, these do not typically account for the indirect effects on AMR due to transmission and thus fail to capture population-level dynamics – they may even yield associations that have the opposite sign from the effects at the population level.^18^

To our knowledge, no study has yet examined the impact of antimicrobial use on susceptibility patterns across multiple antimicrobials (‘phenotypes’)^19^ and multiple pathogens accounting for both individual-level and group-level effects. Under the guiding hypothesis that each antimicrobial exerts selection *for* the phenotypes that are resistant to it, we conducted hierarchical multinomial logistic regression analyses for four major community- and hospital-associated pathogens in the Veterans Affairs Healthcare system (HS) in the United States (U.S.): *S aureus, E coli, K pneumoniae*, and *P aeruginosa*. We included data on antimicrobial susceptibility patterns from individual-level clinical isolates and facility-level antimicrobial prescribing from 138 Veterans Affairs Medical Centres (VAMCs) across all U.S. census regions from 2007-2021. By linking patient-level outcomes to facility-level antimicrobial use, we aimed to investigate how institutional prescribing practices shape resistance patterns observed in patients. The scope of our study - the number of species, diversity of geographic sites, and long period of study - provides a unique opportunity to evaluate this question.

## Methods

### Study design and participants

In this U.S. nationwide retrospective cohort study, we analysed clinical microbiology data from patients admitted to all VA Medical Centers with acute care wards (either medical or surgical wards or intensive care units) from Feb 1, 2007 to Dec 31, 2021. We defined the pre-COVID-19 period as Feb 1, 2007 till Dec 31, 2019 and the COVID-19 period as Jan 1, 2020 till Dec 31, 2021. Patient age and biological sex were determined from birth certificates. Race and ethnicity were self-reported.

This study received ethical approval by the University of Utah Institutional Review Board and the VA Salt Lake City Health Care System Research and Development Office. The study followed the Strengthening the Reporting of Observational Studies in Epidemiology (STROBE) reporting guideline for cohort studies.

### Procedures

Clinical diagnostic cultures were extracted from electronic health records.^20^ Our analysis included only clinically directed cultures, ordered for suspected infection. A 30-day incident isolate was defined as a clinical culture with no prior clinical culture of the same species in the previous 30 days and was used as a proxy for clinical infection incidence. When multiple species were recovered from the same sample or from different samples during the same hospitalisation, each incident isolate was counted separately in the analysis of a given species. Specimens collected more than 3 days post-admission but before discharge were classified as hospital-onset and included in our analysis.

Antimicrobial susceptibility test (AST) results were used to determine resistance to specific antimicrobials. Interpretations, based on the reported minimum inhibitory concentrations, followed CLSI breakpoint revisions and were categorised into susceptible (S), intermediate (I), or resistant (R); intermediate and resistant were grouped as R. We defined (resistance) phenotypes as susceptibility profiles of an isolate to a panel of key antimicrobials selected for their clinical relevance for first- and second-line treatment of the target pathogens (Table 1, appendix pp.7). We referred to isolates susceptible to all three key antimicrobial classes as *susceptible* isolates.

**Table 1.** Descriptive statistics of clinical cultures included in the multinomial logistic regression analysis by organism, 1 Feb, 2007 – 31 Dec, 2021. Patient age was determined from the date of birth. The biological sex of patients was determined from their birth certificates. Race and ethnicity were collected by self-report. Only the most common groups are displayed in this table. Definitions of individual drugs within key antimicrobial classes are provided in the appendix p.4.

|  | Gram-positive cocci | Enterobacterales |  | Gram-negative non-fermenter |
| --- | --- | --- | --- | --- |
|  | *Staphylococcus aureus* | *Escherichia coli* | *Klebsiella pneumoniae* | *Pseudomonas aeruginosa* |
| Key antimicrobial classes | FQL – ASBL – MACR | FQL – 3/4GC – BL/BLI | FQL – 3/4GC – BL/BLI | FQL – BL/BLI – CPM |
| Incident isolates, n | 17,722 | 25,777 | 22,908 | 19,299 |
| Number of facilities | 84 | 100 | 96 | 96 |
| **Sex/Age, n (%)** |  |  |  |  |
| Male | 17,258 (97.4%) | 24,461 (94.9%) | 22,114 (96.5%) | 18,862 (97.7%) |
| Female | 464 (2.6%) | 1,316 (5.1%) | 794 (3.5%) | 437 (2.3%) |
| Median age (IQR), years | 68 (61-77) | 70 (63-79) | 70 (63-78) | 70 (63-78) |
| **Race and ethnicity, n (%)** |  |  |  |  |
| White | 13,085 (73.8%) | 17,525 (68%) | 15,374 (67.1%) | 13,342 (69.1%) |
| Black or African American | 3,381 (19.1%) | 6,513 (25.3%) | 6,081 (26.5%) | 4,650 (24.1%) |
| Hispanic or Latino | 1,636 (9.2%) | 2,102 (8.2%) | 2,354 (10.3%) | 1,978 (10.2%) |
| **Culture sites, n (%)** |  |  |  |  |
| Blood, n (%) | 2,317 (13.1%) | 1,826 (7.1%) | 2,395 (10.5%) | 876 (4.5%) |
| Urine, n (%) | 667 (3.8%) | 14,727 (57.1%) | 10,392 (45.4%) | 7,487 (38.8%) |
| Respiratory, n (%) | 7,840 (44.2%) | 3,758 (14.6%) | 6,450 (28.2%) | 6,995 (36.2%) |
| Other, n (%) | 6,898 (38.9%) | 5,466 (21.2%) | 3,671 (16%) | 3,941 (20.4%) |

\* FQL = fluoroquinolones; ASBL = Anti-Staphylococcal Beta-lactams; MACR = macrolides; 3/4GC = 3<sup>rd</sup> and 4<sup>th</sup> generation cephalosporins; BL/BLI = beta-lactam/beta-lactamase inhibitors; CPM = carbapenems

### Outcomes

Our main outcomes were time trends for incidences of target antimicrobial-pathogen combination phenotypes, facility-level inpatient antimicrobial prescribing rates, and regression coefficients obtained from multinomial logistic regression models. We defined phenotype incidence as the number of incident isolates with a specific susceptibility profile per 1,000 admissions. We defined inpatient antimicrobial prescribing rates as the days of therapy (DOT) per 1,000 patient-days. Multinomial logistic regression coefficients are reported as percentage increase in odds corresponding to one patient-day of use per 100 patient-days in the preceding 14 days when compared to the reference.

### Statistical Analysis

For trends in inpatient antimicrobial prescribing, we included 138 VAMC and performed robust Poisson regression using generalized estimating equations (GEE). Clustering by VAMC was modelled with an autoregressive correlation structure within a facility. We used calendar year as a continuous variable as the main exposure and the number of admissions as offset. We adjusted for major facility characteristics, including census region, facility complexity and rurality, and patient volume, and therefore, accounted for spatial variation and population density (appendix section S2). We estimated the multiplicative average annual percentage change (AAPC) based on the estimated marginalized time trend from our GEE analyses.

To evaluate the relationship between facility-level antimicrobial exposure to resistance phenotypes (i.e., susceptibility profiles to key antimicrobial classes), we fitted mixed-effects multinomial logistic regression models. Facilities were included using a patchwork approach (described below, Table 1). For each pathogen, the outcome was the isolate’s phenotype using the fully susceptible phenotype (S-S-S) as reference. The primary exposure of interest was *recent prescribing*, defined as the proportion of patient-days with antimicrobial prescribing during the 14 days prior to isolate collection for each relevant drug class. To address potential ecological confounding, we adjusted for calendar year, community prevalence (measured as incidence rate of phenotypes collected within 3 days of admission during the previous 90 days), and facility-level characteristics (e.g., census region, facility complexity, facility rurality). Random intercepts for each VA facility were included to capture between-facility heterogeneity. More details and a visual summary are given in the appendix pp.5.

For some antimicrobial classes, such as fluoroquinolones, a substantial proportion of isolates did not have reported AST results, complicating the construction of resistance phenotypes. The incidence and proportion of these missing AST results varied over time, indicating selective reporting.^21^ To address this issue, we applied a threshold-based *patchwork approach* in our main analysis. For each antimicrobial-pathogen combination and each calendar year, we included only those facilities where the proportion of missing AST results did not exceed 30% in each quarter of that year. By excluding facility–year combinations with excessive missing data, we minimized the likelihood that our findings could be skewed by selective reporting. The final dataset is a composite of facilities contributing data in different calendar years. We performed sensitivity analyses to evaluate alternative methods for handling missing AST results (appendix pp.5).

To assess the impact of variation in facility-level antimicrobial prescribing on the ability to recover true regression coefficients, we conducted a simulation analysis assuming the model was correctly specified (the model in the simulation was the same used for fitting). We used a multinomial logistic regression model where the covariates were calendar time and facility-level antibiotic use. We generated three prescribing distributions with the same mean but progressively larger standard deviations, representing low (drug 1), moderate (drug 2), and high variability (drug 3). This analysis examined how variability in prescribing influences bias, confidence interval coverage, the envelope, and the average confidence interval width (appendix pp.22).

## Results

Overall, phenotype incidence trends showed a decline from 2007 to 2021, with only a few notable exceptions (Figure 1, appendix pp.7-24). For *S aureus, K pneumoniae*, and *P aeruginosa* the incidence of isolates resistant to all three key antimicrobial classes (R-R-R) declined from 2007 to 2021. Among these, the incidence of *S aureus* isolates resistant to fluoroquinolones (FQL), Anti-Staphylcoccal Beta-lactams (ASBL), and macrolides (MACR) showed the largest decline (AAPC=-9.5%; 95%CI: −11.6%, −7.3%; p-value<0.0001). In contrast, the incidence of *E coli* isolates resistant to FQL, third and fourth generation cephalosporins (3/4GC), and β-lactam/β-lactamase inhibitors (BL/BLIs) increased from 2007 to 2021 (AAPC=3.4%; 95%CI: 0.5%, 6.5%; p-value=0.0204). In addition, the incidence of 3/4GC-resistant *E coli* isolates increased. In particular, the incidence of R-R-S increased with an AAPC of 4.9% (95%CI: 0.6%, 9.4%; p-value=0.0264). The incidence of *E coli* phenotypes susceptible to 3/4GC (R-**S**-R, R-**S**-S, S-**S**-R, S-**S**-S) declined. The largest decline was estimated for FQL-resistant *E coli* susceptible to both 3GC and BL/BLI (R-S-S; AAPC:-8%, 95%CI: −11.3%, −4.6%; p-value<0.0001). The incidence of susceptible isolates (S-S-S) declined from 2007 till 2021 for all target pathogens, with larger declines estimated for Enterobacterales species *E coli* (AAPC:-5.1%, 95%CI: −6.8%, −3.5%; p-value<0.0001) and *K pneumoniae* (AAPC:-5.5%, 95%CI: −7.1%, −4%; p-value<0.0001) than for others.

**Figure 1.**
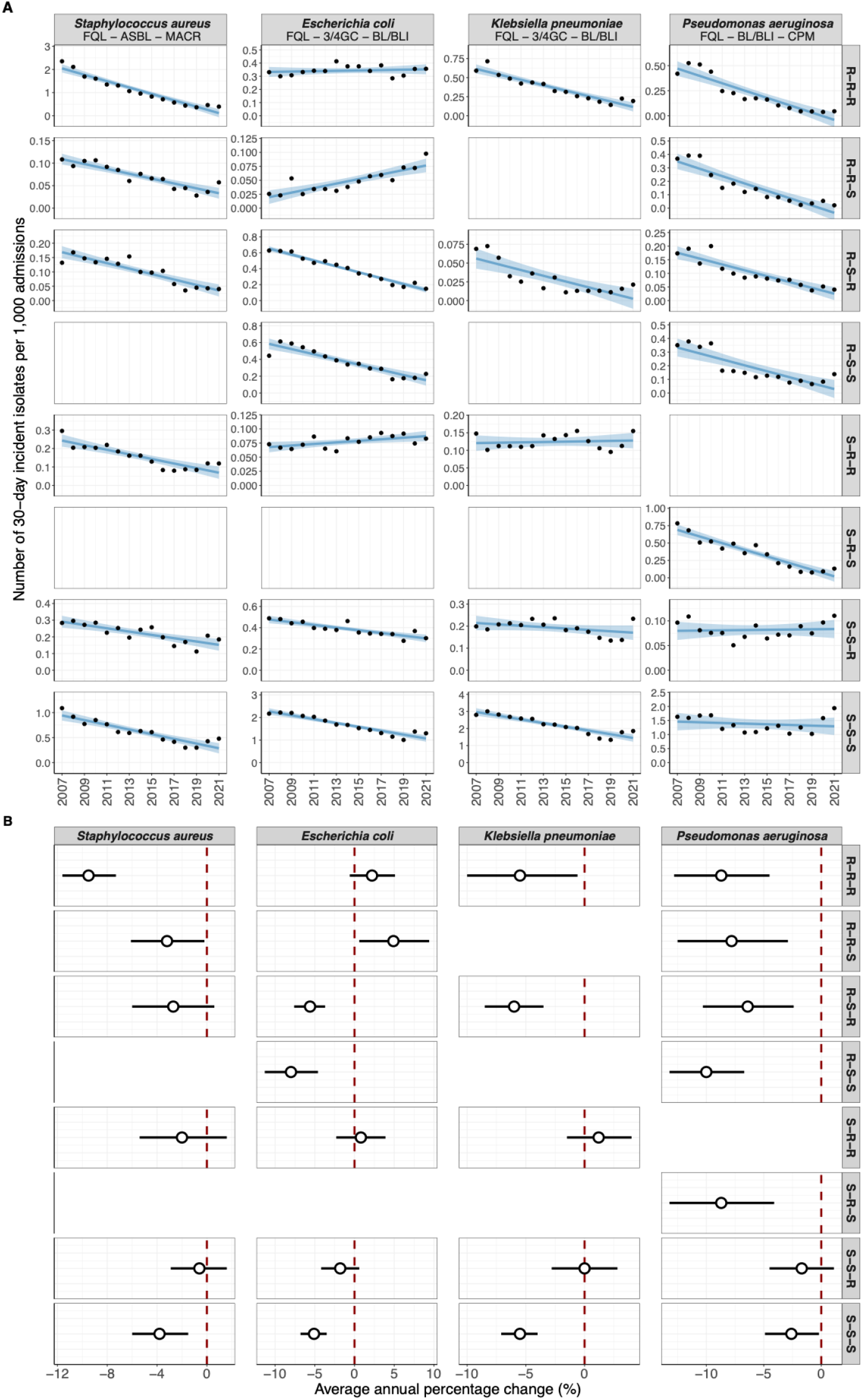
Trends in antimicrobial resistance phenotypes among hospital-onset isolates of four target organisms in the Veterans Affairs Healthcare Administration, February 1, 2007–December 31, 2021. Patchwork data were used for analyses, as described in the appendix (p.□29). (A) Points show the incidence of hospital-onset isolates with the respective phenotype. Lines represent linear regression fits with shaded 95% confidence intervals. (B) Average annual percentage change (AAPC) estimates from time trend analyses using generalized estimating equations for phenotypic incidence. Interactions with the COVID-19 period were evaluated; separate estimates are shown if the interaction term was statistically significant (p < 0.05, Bonferroni-corrected). Positive AAPC values indicate increasing trends, and negative values indicate decreasing trends. FQL = fluoroquinolones; ASBL = Anti-Staphylococcal Beta-lactams; MACR = macrolides; 3/4GC = 3rd and 4th generation cephalosporins; BL/BLI = beta-lactam/beta-lactamase inhibitors; CPM = carbapenems. AAPC = average annual percentage change; R = resistant or intermediate; S = susceptible.

Inpatient prescribing declined for most antimicrobial classes (Figure 2, appendix p.27). The largest reduction was estimated for FQL from 2007 to 2022 (AAPC=-8.4%, 95%CI: −9.1%, −7.7%; p-value<0.0001). Carbapenem prescribing rates increased between 2007 and 2011 (AAPC=5.3%; 95%CI: 3.1%, 7.6%; p-value<0.0001) and subsequently declined between 2012 and 2022 (AAPC=-3.7%, 95%CI: −6.2%, −1.1%; p-value=0.0062). BL/BLI prescribing rates stayed stable from 2007 till 2014 (AAPC: −0.1%, 95%CI: −0.8%, 0.6%; p-value=0.7993). After a nationwide shortage of piperacillin/tazobactam in 2015, BL/BLI prescribing rates declined (AAPC=-2.3%, 95%CI: −3.9%, −0.7%; p-value=0.0046). Prescribing rates increased for 3/4GC from 2007 to 2019 (AAPC=3.1%, 95%CI: 2.1%, 4.1%; p-value<0.0001) but subsequently declined during the COVID-19 pandemic years (2020-2022; AAPC=-2.4%, 95%CI: −5.2%, −0.5%; p-value=0.1071), although the confidence interval included the null. Prescribing of ASBL and MACR declined modestly from 2007 to 2019 (AAPC=–1.0%, 95%CI: –1.7%, −0.2%; p-value=0.0103, and AAPC=-0.1%, 95% CI: −1.0%, 0.9%; p=0.8488, respectively). During the COVID-19 pandemic (2020–2022), ASBL prescribing increased (AAPC=4.6%, 95%CI: 1.5%, 7.8%; p-value=0.0038) while MACR prescribing fell sharply (AAPC=–17.9%, 95%CI: −21.0%, −14.7%; p-value<0.0001).

**Figure 2.**
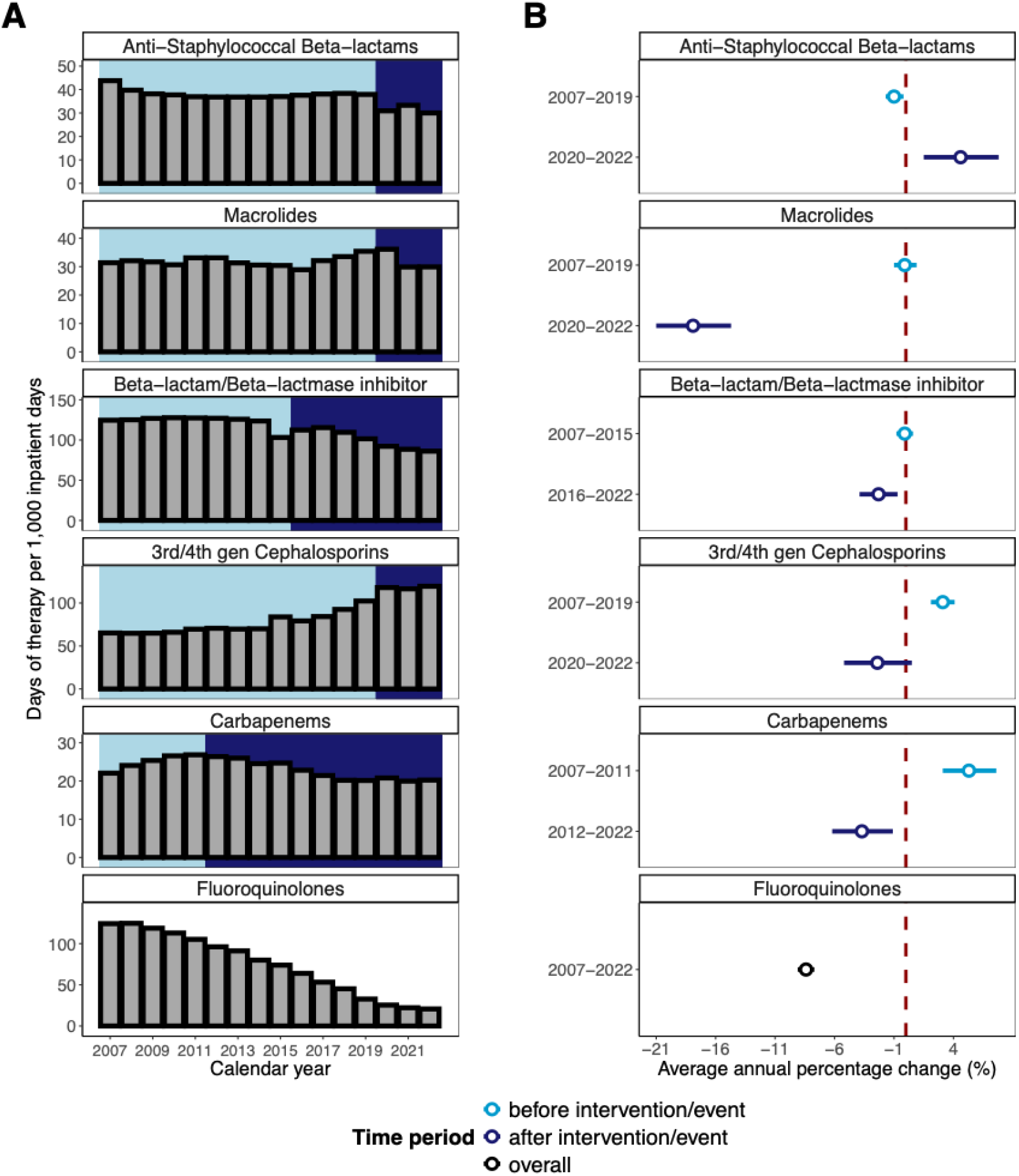
Trends in antimicrobial prescribing in the Veterans Affairs Healthcare Administration, February 1, 2007–March 31, 2022. (A) Bar plots show overall antimicrobial prescribing rates, expressed as days of therapy per 1,000 patient-days, in 138 VA medical centers for key antimicrobial classes. Shaded regions indicate time periods corresponding to those analysed in panel B. (B) Average annual percentage change (AAPC) estimates from time-trend analyses using generalized estimating equations for the antimicrobial classes in panel A. Positive AAPC values indicate increasing trends; negative values indicate decreasing trends. For anti-staphylococcal β-lactams, macrolides, third-generation cephalosporins, and fluoroquinolones, interactions with the COVID-19 period were evaluated, and separate trend estimates are presented if the interaction term was statistically significant (overall p < 0.05, Bonferroni-corrected). For β-lactam/β-lactamase inhibitors and carbapenems, interactions with 2015 drug shortages and 2011 antimicrobial stewardship initiatives, respectively, were similarly evaluated. “Before” and “after” denote the periods preceding and following these breakpoints, as shown on the y-axis. Further details are provided in the appendix (pp.□25–28).

Although unadjusted single antimicrobial-single pathogen associations between antimicrobial use and resistance were generally weak (appendix p.25), our multinomial logistic regression analyses revealed that recent FQL prescribing consistently increased the relative odds of FQL-resistant phenotypes across all four target pathogens (Figure 3). Specifically, point estimates for recent FQL prescribing indicated higher odds of isolating FQL-resistant phenotypes (compared to the reference S-S-S), with confidence intervals excluding the null effect in 11 out of 13 cases. Among *S aureus* isolates, the strongest association was estimated for FQL‐resistant MRSA (R-R-S) isolates, for which one additional treatment day (per 100 patient-days) increased the odds of isolating a R-R-S phenotype (FQL-resistant, MACR-susceptible MRSA) by 4.6% (95%CI: 0.6%, 7.7%, p-value=0.0032). Macrolide prescribing was linked to increasing the odds of isolating FQL-susceptible, MACR-resistant MRSA (S-R-R) by 4.5% (95% CI: 0.4%, 8.8%, p-value=0.0302). In contrast, ASBL prescribing did not increase the odds of ASBL resistance for any of the studied phenotypes. Paradoxically, each additional treatment day (per 100 patient-days) was associated with a 4.6% reduction in the odds of fluoroquinolone-resistant MRSA (95%CI: −10.1%, 1.3%, p-value=0.1257), although the wide confidence interval included the null effect.

**Figure 3.**
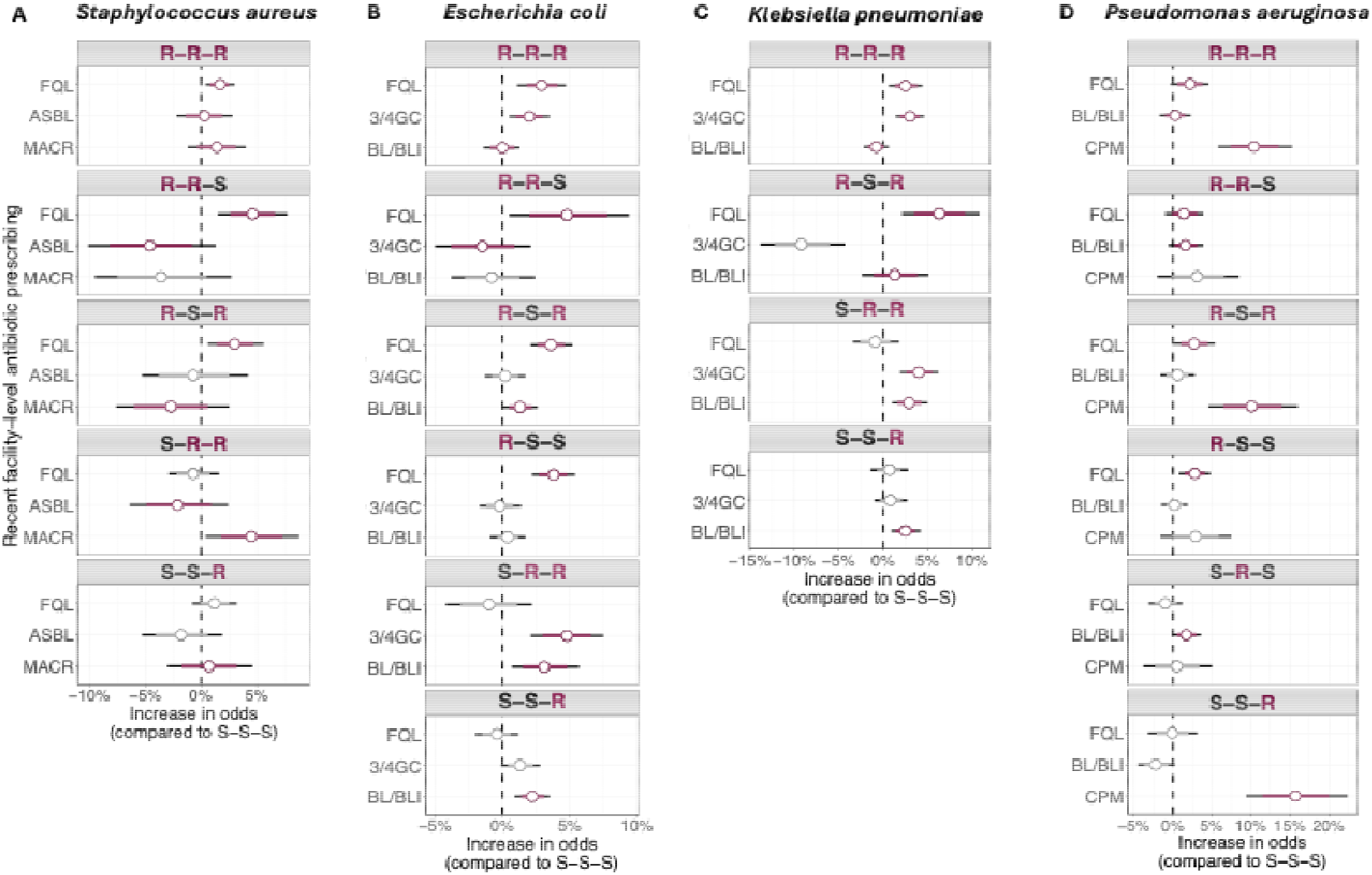
Effect of recent antimicrobial prescribing on resistance patterns in four bacterial pathogens. Patchwork data was used for the analysis and the approach was described in the appendix p.28. Multilevel multinomial logistic regression coefficients are shown as the percentage change in the odds of each resistance phenotype for every additional treatment day (per 100 patient-days) of exposure to the specified antimicrobial class in the preceding 14 days (per 1,000 patient-days), compared with isolates fully susceptible to all key classes (S-S-S). Points denote medians; thick bars, 80 % confidence intervals; thin bars, 95 % confidence intervals. Coefficients highlighted in dark red correspond to antimicrobial-pathogen combinations for which an effect of prescribing on resistance was hypothesised. Antibiotic classes for each pathogen: (A) *S aureus:* fluoroquinolones, anti-staphylococcal beta-lactams, macrolides. (B) *E coli:* fluoroquinolones, 3^rd^ and 4^th^ generation cephalosporins, beta-lactam/beta-lactamase inhibitors. (C) *K pneumoniae:* fluoroquinolones, 3^rd^ and 4^th^ generation cephalosporins, beta-lactam/beta-lactamase inhibitors. (D) *P aeruginosa:* fluoroquinolones, beta-lactam/beta-lactamase inhibitors, carbapenems. FQL = fluoroquinolones, ASBL = anti-staphylococcal beta-lactams, MACR = macrolides, 3/4GC = 3^rd^ and 4^th^ generation cephalosporins, BL/BLI = beta-lactam/beta-lactamase inhibitor, CPM = carbapenems.

For *E coli*, the largest effect of FQL prescribing was estimated for isolates resistant to FQL but susceptible to 3GC and BL/BLIs (R-S-S, Figure 3B) and for isolates resistant to both FQL and BL/BLI (R-S-R, Figure 3B) for which the relative odds were increased by 3.6% (95%CI: 2%, 5.2%, p-value<0.0001) and 3.6% (95%CI: 2.0%, 5.1%, p-value<0.0001) for each additional day of FQL treatment (per 100 patient-days), respectively. In *K pneumoniae*, one additional treatment day of FQL (per 100 patient-days) increased the odds of isolates resistant to both FQL and BL/BLIs (R-S-R) by 6.3% (95%CI: 1.8%, 11%, p-value=0.0057). For *P aeruginosa*, one additional FQL treatment day increased the odds by 2.8% (95%CI: 0.8%, 4.8%, p-value=0.0051) for phenotypes resistant to only FQL (R-S-S) and by 2.2% (95%CI: −0.3%, 4.6%, p-value=0.0798) for phenotypes resistant to all three key antimicrobial classes.

For both Enterobacterales species, recent 3/4GC and BL/BLI prescribing was generally associated with higher relative odds of resistance to these drugs (appendix pp.11-15), with strongest effects for phenotypes resistant to both 3/4GC and BL/BLI. For example, each additional 3/4GC treatment day (per 100 patient-days) increased the odds of *E coli* S-R-R isolates by 4.8% (95%CI: 2.1%, 7.5%, p-value=0.0003) and of R-R-R isolates by 2.0% (95%CI: 0.5%, 3.5%, p-value=0.0102). No association was found between 3/4GC prescribing and R⍰R⍰S isolates or between BL/BLI prescribing and isolates resistant to all three key antimicrobial classes (R-R-R). In sensitivity analyses, 4GC prescribing appeared to be the primary driver of the observed associations, showing a link with phenotypes resistant to 4GCs but not with those susceptible to 4GCs and resistant to 3GCs (appendix p.15). In contrast, 3GC prescribing was not associated with 3GC-resistant phenotypes.

Among the antibiotic exposures evaluated for *P aeruginosa*, recent antipseudomonal CPM prescribing demonstrated the strongest association with corresponding resistance phenotypes. One additional CPM treatment day increased the odds by 15.7% (95%CI: 9.4%, 22.4%, p-value<0.0001) for CPM-resistant *P aeruginosa* isolates susceptible to both FQL and BL/BLIs (S-S-R), by 10.5% (95%CI: 5.7%, 15.3%, p-value<0.0001) for isolates resistant to FQL, BL/BLIs and CPM (R-R-R), and by 10.4% (95%CI: 5.8%, 15.3%, p-value<0.0001) for isolates resistant to FQLs and CPMs (R-S-R). Recent CPM prescribing showed larger associations with resistant phenotypes than recent FQL prescribing (Figure 3D).

To assess robustness, we used a rescaled exposure metric to reflect a 1% increase in average antimicrobial prescribing volume. While the results remained broadly consistent, the difference in effect size between CPM and FQL prescribing diminished (appendix p.24).

For most pathogen-phenotype combinations, the importation of phenotypes by community-onset isolates was significantly associated with an increase in odds of isolates with that specific phenotype (appendix section S4).

To examine how variability in prescribing patterns influences coefficient estimation, we conducted a simulation analysis with a correctly specified multilevel multinomial logistic regression model (Figure 4; appendix p.30). Lower variability (Drug 1) had little effect on the median bias; however, it markedly increased its variance (Figure 4A). Lower variability also broadened both the range of the estimated coefficients (envelope) and the mean widths of their 95% confidence intervals (Figure 4C), reflecting greater statistical uncertainty. The coverage, the proportion of 95% confidence intervals that contained the true coefficient, remained stable across the scenarios (Figure 4B).

**Figure 4.**
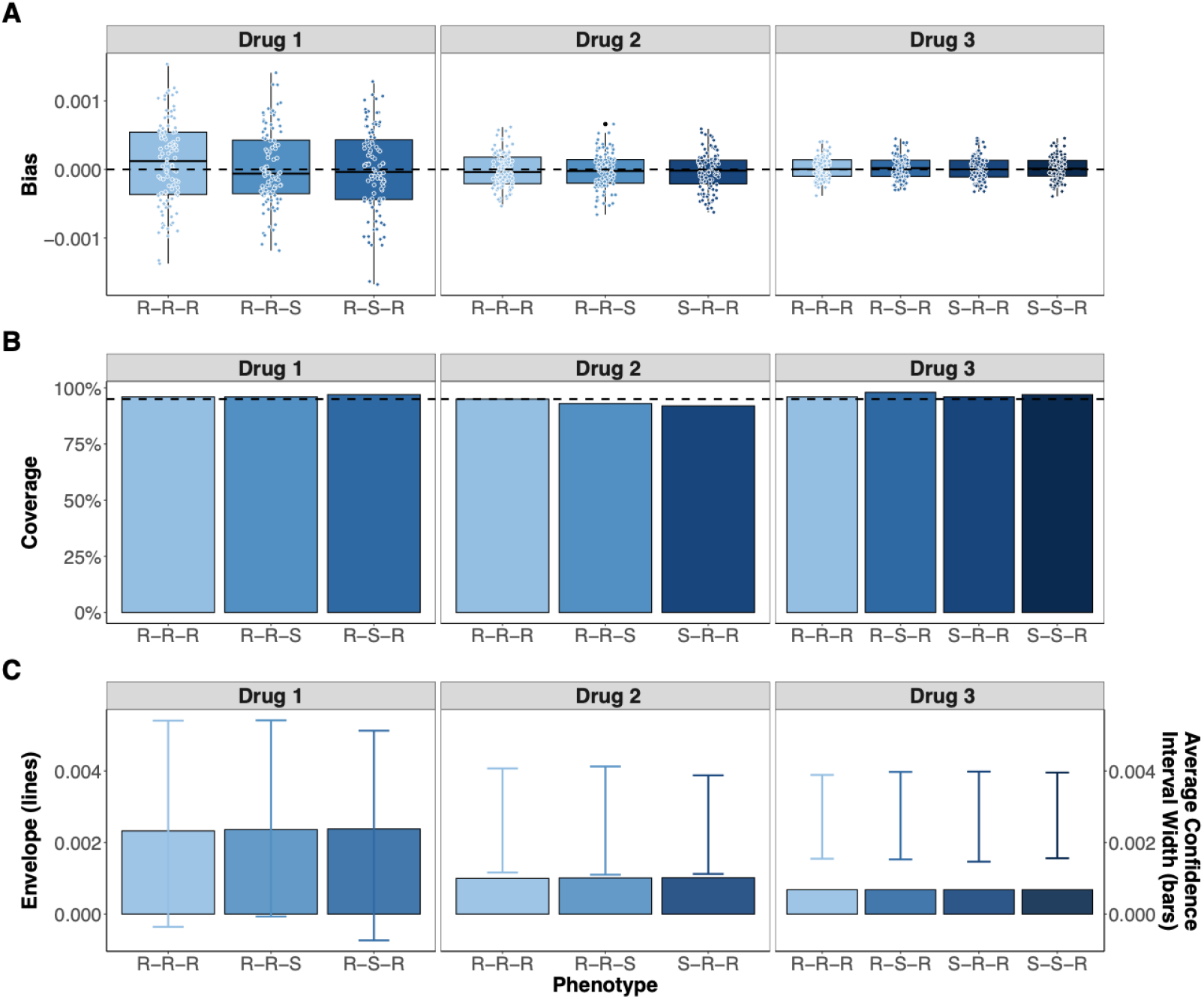
Impact of variability in facility-level antimicrobial prescribing on regression coefficient estimation on bias, coverage, envelope and confidence interval precision. Three prescribing distributions of drug classes were simulated with low (Drug 1), medium (Drug 2), and large variance (Drug 3) based on log-normal distribution with parameters described in the appendix (section S5, p.27-28). The results were summarized across 100 simulated data sets. (A) Boxplot and distribution of bias in regression coefficient estimates, defined as the difference between the estimate and the true coefficient. (B) Bar plots of coverage of 95% confidence intervals. This panel illustrates the proportion of simulation runs in which the true regression coefficient is captured by the estimated 95% confidence intervals. (C) Envelope (lines, left y-axis) and Average Confidence Interval Width (bar plots, right y-axis). The envelope (range) of estimated regression coefficients is shown as line intervals (left y-axis) and the average width of the 95% confidence intervals is represented by bar plots (right y-axis).

## Discussion

In this nationwide study, we observed notable shifts in phenotypic incidence trends among four major pathogens in the VAHS between 2007-2021, in parallel with changes in antimicrobial prescribing. By decomposing resistance into distinct phenotypes, we identified patterns suggestive of co-selection. Multinomial logistic regression enabled us to disentangle the individual effects of specific antimicrobials on resistance patterns – an advantage over traditional single pathogen-single antimicrobial approaches. Facility-level prescribing was significantly associated with resistance at the individual level across most pathogen-antimicrobial combinations. These findings provide a framework for anticipating how shifts in antimicrobial use may shape future resistance dynamics.

A particularly notable finding was the broad decline in FQL resistance across all four pathogens and eleven phenotypes, except for *E coli* resistant to both FQL and 3GC. These trends were supported by significant associations between facility-level FQL prescribing and FQL resistance in all target pathogens. In particular, our results suggest that MRSA incidence may be driven by FQL and MACR prescribing rather than anti-staphylococcal beta-lactams, echoing previous findings that linked FQL but not beta-lactam use to MRSA colonization.^17,22^ Several studies linked FQL restriction to a reduction in MRSA isolation rates,^23,24^ and a 10-year time-series analysis showed MRSA rates increased upon reintroduction of FQLs.^25^ This pattern, together with evidence that FQL promote the loss of MSSA colonization more than other anti-MSSA agents,^26^ suggests that fluoroquinolones may promote MRSA selection through ecological disruption, eradicating MSSA strains and creating niches for resistant strains to proliferate.^27^ The lack of association between anti-staphylococcal beta-lactams and MRSA incidence in our study may reflect the distinct evolutionary pathway of methicillin resistance. Historically, penicillin use – not methicillin – has been implicated as the initial driver of *mecA* acquisition in *S aureus*.^28^

Prescribing of FQL, CPMs and BL/BLs were associated with subsequent resistance in *P aeruginosa*, consistent with prior reports linking the use of these antibiotics with MDR and extensively resistant (XDR) *P aeruginosa* infections.^29,30^ Additionally, CPM restrictions have been associated with reduced CPM resistance incidence.^31^ We observed declining incidences of all studied resistant phenotypes, consistent with previously described reductions in hospital-associated infections in the VAHS and of MDR/XDR *P aeruginosa* in other regions.^21^ These trends paralleled reductions in FQL, CPM, and BL/BLI prescribing in our study. Further research is needed to disentangle whether these declines are fully attributable to reductions in antibiotic use or reflect broader hospital-wide interventions such as infection control measures.

The persistence of FQL resistance in *E coli* isolates, despite declining FQL prescribing, highlights the complexity of AMR dynamics. Over the study period, 3/4GC prescribing increased, potentially as a substitute for FQL. Higher recent facility-level exposure to 3/4GC was generally associated with increased odds of phenotypes resistant to 3/4GCs. Sensitivity analyses suggested that this effect was primarily driven by 4GC exposure rather than 3GC. Further research is needed to determine whether this is due to stronger selective pressure exerted by 4GCs or other factors. Additionally, while the incidence of *E coli* isolates resistant to 3/4GCs increased, the incidence of 3GC-susceptible *E coli* phenotypes declined, suggesting that 3/4GC use may help maintain or even drive FQL and BL/BLI resistance in MDR *E coli*. These findings indicate that reductions in prescribing for one antimicrobial class may inadvertently drive resistance through increased prescribing of another, potentially leading to co-resistance. Continued surveillance and studies are essential to identify and further characterize these relationships. Given that once resistance emerges, large reductions in consumption are required to meaningfully reduce resistance,^32^ early and multifaceted stewardship and infection control interventions are essential to controlling AMR.

Our study has several limitations. First, although our model explicitly accounts for prior antimicrobial exposure, community prevalence, and facility-level characteristics, residual confounding, including reverse causation, cannot be excluded. For instance, facilities with a high prevalence of MRSA may deliberately restrict agents linked to MRSA selection, which could paradoxically associate higher prescribing of anti⍰staphylococcal beta⍰lactams with lower MRSA prevalence. The 14⍰day lag in prescribing we incorporated to mitigate reverse causality may be insufficient to capture such dynamics. Second, only inpatient prescriptions were analysed. For many pathogens, community-based prescribing may contribute significantly to selection pressure and subsequent hospital importation of resistant organisms. Third, prescribing data served as a surrogate for actual antimicrobial consumption since information on patient-level adherence was not available. Fourth, we classified isolates as hospital-onset if they were collected at least 72 hours after admission, which can misclassify some community-acquired infections. Fifth, our simulation study indicates that precise estimation of regression coefficients requires substantial variability in corresponding prescribing. Limited variability for certain antibiotic classes may have inflated uncertainty and contributed to some of our null findings. Sixth, while phenotypic resistance patterns can approximate bacterial lineages, integrating whole⍰genome sequencing with phenotypic data in future work would more clearly elucidate links between antimicrobial use and microbial ecology.^33^ Finally, the study population comprised US veterans receiving care in VAMCs. This cohort differs from the general population in demographics, comorbidities, exposures, and health⍰care utilisation. Nonetheless, our dataset spans over 100 hospitals of varying complexity across diverse U.S. regions over more than 15 years, supporting the general relevance of our findings.

Overall, our findings highlight the complexity of antimicrobial selection and its variable impact on resistance dynamics across drug classes, pathogens, and underlying resistance mechanisms. Judicious prescribing is essential: reducing use of one drug class may inadvertently increase reliance on – and resistance to – another. Effective antimicrobial stewardship must take a comprehensive approach, targeting multiple drug classes simultaneously. Such coordinated strategies, guided by surveillance and timely interventions, are crucial to limiting the spread of multidrug resistance and preserving the effectiveness of current therapies.

## Data Availability

All data produced in the present study are available upon reasonable request to the authors.

## Data sharing

Individual-level patient data cannot be provided due to Veteran Affairs privacy practices. Artificial data that represents the original data will be provided along with a data dictionary that defines each field in the dataset and supporting documentation (statistical and analytic code) are published on Github (https://github.com/tm-pham/mind_aim2-2_AMR).

## Acknowledgments

This work was supported by the US Centers for Disease Control and Prevention (Modeling and Simulation to Support Epidemiological Decision-Making in Healthcare Settings, Modeling Infectious Diseases in Healthcare Program; U01CK000585). MS was supported by the Agency for Healthcare Research and Quality (R01HS025175) and the Department of Veterans Affairs (1I50HX002731–01). We thank the Department of Veterans Affairs for the supported infrastructure and data resources. The views expressed in this article are those of the authors and do not necessarily represent the position or policy of the Department of Veterans Affairs, the US Centers for Disease Control and Prevention, the US Government, or any of the affiliated institutions.

## Declaration of generative AI and AI-assisted technologies in the writing process

During the preparation of this work the authors used ChatGPT to improve clarity of the written manuscript. After using this tool/service, the authors reviewed and edited the content as needed and take full responsibility for the content of the publication.

