## Supplement for "Antimicrobial selection for resistance in four major pathogens in the US Veterans Affairs Healthcare System, 2007-2021"

### S1 **Data**

Data used in this analysis were previously described in Pham et al (2024).^1^ In short, the main source for the data of this study was the Veterans Affairs Corporate Data Warehouse (CDW), which is a national repository that includes clinical and administrative data from the VHA. These data are updated on a continual basis and additional information can be found here: <https://www.hsrd.research.va.gov/for_researchers/cdw.cfm>.

Two large datasets were generated from the Veterans Health administration’s (VHA’s) electronic health records with over 15 years of data from all patients admitted to acute care wards (either medical/surgical wards or ICUs) from Veterans Affairs Medical Centres (VAMCs) between January 1, 2007 and March 31, 2022 for the analyses of this study.

The first dataset represents a hospital-day level dataset for all acute care hospitalizations and any administered antibiotics on those days and contains over 50,000,000 hospital days. It includes information on patient demographics, comorbidities, mortality, readmission information, risk factors, and primary diagnosis categories. We used days of therapy divided by occupied bed days (bed days of care) as the primary measure of antibiotic use, in concordance with National Health Safety Network (NHSN) guidelines for electronic reporting of antibiotic utilization data.

The second dataset includes all microbiology cultures of the four target organisms (*Staphylococcus aureus, Escherichia coli, Klebsiella pneumoniae, Pseudomonas aeruginosa*) and their antibiotic susceptibility patterns. For our analysis, we considered only positive clinical cultures and removed any surveillance cultures. A descriptive overview of this dataset is given in Table S1. These clinical cultures were obtained only when infection is suspected in symptomatic individuals. The systemic antibiotics were broken out by specific antibiotics (Table S2) and antibiotic (sub)class, and by Standardized Antimicrobial Administration Ratio (SAAR) antimicrobial category. The antibiotic susceptibility test (AST) results for the key antibiotics/antibiotic classes (methicillin, fluoroquinolone, 3rd generation cephalosporins, carbapenem, beta-lactam/beta-lactamase inhibitors) were determined by reported minimum inhibitory concentrations (MIC) for specific antibiotics or by tests of specific genetic determinants (Table S2). Interpretation of AST results adhered to breakpoint revisions by the Clinical and Laboratory Standards Institute. Key antibiotics and antibiotic classes were chosen based on clinical relevance, i.e., first-line and second-line treatment of the target pathogens.

We excluded facilities from the study if they did not provide acute care or if they did not report data to the VA’s facility complexity assessment (e.g., level and type of care provided) in all eligible years during the study period.

Table S1. Descriptive statistics of clinical cultures in the Veterans Affairs Healthcare System by organism, February 1, 2007 – December 31, 2021. Patient age was determined from the date of birth. The biological sex of patients was determined from their birth certificates. Race and ethnicity were collected by self-report. Only the most common groups are displayed in this table.

|  | **Gram-positive cocci** | **Enterobacterales** | | **Gram-negative non-fermenter** |
| --- | --- | --- | --- | --- |
|  | ***Staphylococcus aureus*** | ***Escherichia coli*** | ***Klebsiella pneumoniae*** | ***Pseudomonas aeruginosa*** |
| Incident isolates, n | 247,329 | 222,098 | 13,7923 | 130,496 |
| Number of facilities | 138 | 139 | 137 | 138 |
| **Sex/Age, n (%)** | | | | |
| Male, n (%) | 239040 (96.6%) | 204271 (92%) | 131229 (95.1%) | 127157 (97.4%) |
| Female, n (%) | 8288 (3.4%) | 17827 (8%) | 6693 (4.9%) | 3339 (2.6%) |
| Median age (IQR), years | 66 (59-75) | 71 (63-81) | 70 (63-79) | 71 (63-80) |
| **Race and ethnicity, n (%)** | | | | |
| White, n (%) | 188964 (76.4%) | 156882 (70.6%) | 96760 (70.2%) | 95636 (73.3%) |
| Black or African American, n (%) | 42986 (17.4%) | 50674 (22.8%) | 32511 (23.6%) | 26771 (20.5%) |
| Hispanic or Latino, n (%) | 14644 (5.9%) | 16800 (7.6%) | 10700 (7.8%) | 9218 (7.1%) |
| **Culture sites, n (%)** | | | | |
| Blood, n (%) | 34496 (13.9%) | 24491 (11%) | 15212 (11%) | 6068 (4.6%) |
| Urine, n (%) | 28145 (11.4%) | 148361 (66.8%) | 78072 (56.6%) | 56488 (43.3%) |
| Respiratory, n (%) | 48954 (19.8%) | 12780 (5.8%) | 20171 (14.6%) | 34172 (26.2%) |
| Other, n (%) | 135734 (54.9%) | 36466 (16.4%) | 24468 (17.7%) | 33768 (25.9%) |

**Table S2. Overview of specific antibiotics and other genetic determinants to determine antibiotic susceptibility test results in key antibiotic classes.**

| **Anti-Staphylococcal beta-lactams (ASBL)** | **Fluoroquinolones (FQL)** | **3^rd^ / 4^th^ generation cephalosporins (3/4GC)** | **Beta-lactam/**  **Beta-lactamase inhibitors (BL/BLI)** | **Carbapenems**  **(CPM)** |
| --- | --- | --- | --- | --- |
| Cefazolin | Ciprofloxacin | Cefoperazone (3GC) | Amoxicillin/ clavulanate | bla(KPC) gene (test for presence using molecular method) |
| Cefoxitin | Gatifloxacin | Cefotaxime (3GC) | Ampicillin/sulbactam | Carbapenemase gene (by nucleic acid amplification with probe detection) |
| mecA gene | Gemifloxacin | Cefpodoxime (3GC) | Ceftazidim/avibactam | Hodge test |
| Methicillin | Levofloxacin | Ceftazidime (3GC) | Ceftolozane/tazobactam | Imipenem |
| Nafcillin | Lomefloxacin | Ceftizoxime (3GC) | Piperacillin/tazobactam | Meropenem |
| Oxacillin | Moxifloxacin | Ceftriaxone (3GC) | Imipenem/cilastatin/relebactam | Doripenem |
| Dicloxacillin | Norfloxacin | Moxalactam (3GC) | Meropenem/vaborvactam | Ertapenem |
|  | Ofloxacin | Cefepime (4GC) | Ticarcillin/clavulanate |  |

### S2 **Statistical analysis**

#### S2.1 Time trend analyses

We performed time trend analyses for antibiotic prescribing and phenotype incidence using generalized estimating equations (GEE) similar to our methods in Pham et al (2024).^1,2^

#### S2.2 Multinomial logistic regression for use-resistance relationship

We used a mixed-effects multinomial logistic regression to evaluate the association between facility-level antibiotic use and resistance patterns (Figure S1). For each pathogen, the outcome was the phenotype category of each clinical isolate, reflecting a sentinel of facility-level resistance phenotypes. The primary exposure was the proportion of facility-level antibiotic days of therapy in the 14 days prior to the isolate’s collection date. To address potential ecological confounding, we adjusted for calendar time, community prevalence (measured by the incidence rate of resistance phenotypes collected within 3 days of admission in the preceding 90 days), and facility characteristics including census region, facility complexity, and rurality. Given the ecological nature of the analysis, we adjusted only for facility-level confounders. We incorporated facility-level random intercepts to account for within-facility correlations over time, allowing each facility to have its own baseline distribution of phenotype categories while estimating the overall association between facility-level antibiotic use and resistance outcomes.

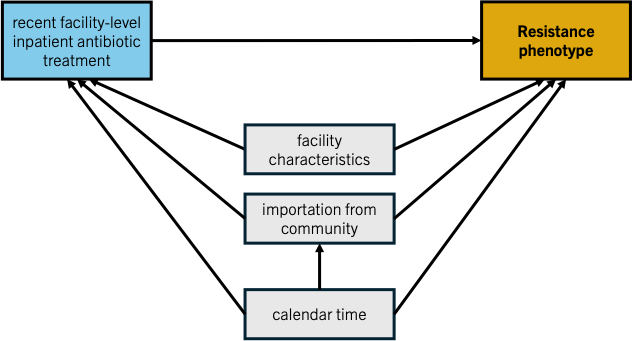

Figure S1. Directed acyclic graph (DAG) illustrating the assumed relationships among covariates, the main exposure, and the outcome for the multinomial logistic regression analysis. The outcome variable (yellow) is the resistance phenotype of each clinical isolate. The primary exposure (blue) is recent facility-level inpatient antibiotic treatment, measured as the proportion of treatment days for a given antimicrobial in the 14 days preceding the isolate’s collection date. Facility-level characteristics (census region, facility complexity, facility rurality, patient composition), importations from the community (prevalence of resistance phenotypes among community-onset isolates in past 3 months), and calendar time (grey) were included as covariates to control for potential confounding.

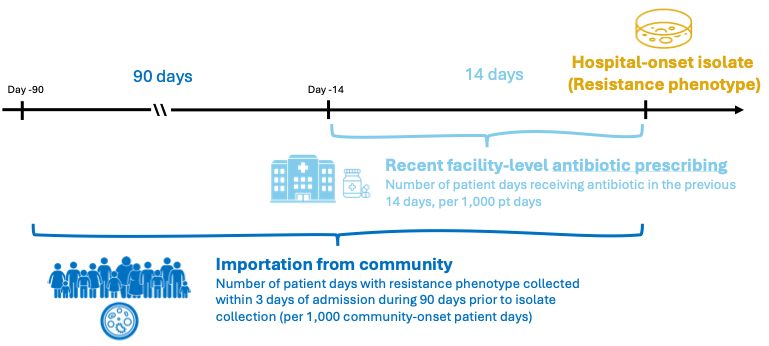

Figure S2. Overview of key exposure and confounder variables of multinomial logistic regression. Visual summary of the temporal structure used to define the main exposure and confounder variables in the multinomial logistic regression model. The outcome was the resistance phenotype of hospital-onset isolates, defined as those collected more than 3 days after admission. The main exposure, recent facility-level antibiotic prescribing, was calculated as the proportion of patient-days with antibiotic use during the 14 days prior to isolate collection. To account for importation of resistance from the community, we included a measure of community prevalence, defined as the number of patient-days with a resistance phenotype collected within 3 days of admission during the preceding 90 days (per 1,000 community-onset patient-days).

**Outcome:**

- Phenotype category $Y_{i}, i=0, 1, \ldots, K-1$ ($K$ categories)
- Reference category: $Y_{0}$

**Random effects**: $u_{k, sta6a}$ (random intercept for each facility)

**Fixed effects:**

Covariates: $X_{pi}, p=1, \ldots, P, i=0, 1, \ldots, K-1$ ($P$ distinct covariates)

Exposure:

- Inpatient facility-level antibiotic exposure (numeric):
  - Facility-level inpatient antibiotic use in the previous 14 days (Number of prescriptions per 100 patient days) for each target antibiotic/antibiotic class

Adjustment variables:

- Temporal
  - Calendar year (numeric)
  - Day of the week (categorical)
  - Month of the year (categorical)
- Importation from community (numeric)
  - Prevalence of phenotype among community-onset isolates in past 3 months (per phenotype category: Number of community-onset isolates divided by total number of isolates)
- Other facility-level variables
  - Census region (South – reference, West, Midwest, Northeast, Puerto Rico and outlying areas)
  - Facility rurality category
    - Urban (reference)
    - Rural
  - Facility complexity level (1a - reference, 1b, 1c, 2, 3)
    - Facility with no assigned complexity level were excluded
  - Patient composition
    - Median length of stay in facility
    - Median Charlson Comorbidity index in facility
    - Median age in facility
    - Percentage of female patients

We used the following reference categories in the analysis:

Table S3. Reference categories in multinomial logistic regression analysis for four target organisms.

| **Organism** | **Antibiogram reference category** |
| --- | --- |
| *Staphylococcus aureus* | S-S-S |
| *Klebsiella pneumoniae* | S-S-S |
| *Escherichia coli* | S-S-S |
| *Pseudomonas aeruginosa* | S-S-S |

The $K-1$regression equations are as follows:

$\log\frac{P\left( Y_{i}=k \right)}{P\left( Y_{i}=0 \right)}=\beta_{0k}+u_{k, sta6a}+\sum_{p=1}^{P} \beta_{pk}X_{pi}$

### S3 **Additional results**

#### S3.1 *Staphylococcus aureus*

**Table S3**. **Most common phenotypes for *Staphylococcus aureus* clinical 30-day incident isolates in the patchwork dataset, from Feb 1, 2007 to Dec 31, 2021.** The phenotype in bold was used as the reference category (S-S-S) in the multinomial logistic regression analysis.

|  | **Fluoroquinolones** | **Anti-Staphylococcal beta-lactams** | **Macrolides** | **Count** | **Percentage** | **Cumulative percentage** | **Abbreviation** |
| --- | --- | --- | --- | --- | --- | --- | --- |
| 1 | R | R | R | 8384 | 39.5 | 39.5 | R-R-R |
| **2** | **S** | **S** | **S** | **4776** | **22.5** | **61.9** | **S-S-S** |
| 3 | S | S | R | 1732 | 8.2 | 70.1 | S-S-R |
| 4 | S | R | R | 1207 | 5.7 | 75.8 | S-R-R |
| 5 | R | S | R | 805 | 3.8 | 79.5 | R-S-R |
| 6 | R | R | S | 555 | 2.6 | 82.2 | R-R-S |

Table S4. Time trend estimates for *S aureus* phenotype incidence, from Feb 1, 2007 to Dec 31, 2021. Time trend estimates represent average annual percentage change (AAPC) and were obtained using generalized estimating equations. This table corresponds to Figure 2B in the main text.

| **Phenotype** | **Time period** | **Time trend estimate** | **95% confidence interval** | | **p-value** |
| --- | --- | --- | --- | --- | --- |
|  |  |  | **Lower bound** | **Upper bound** |  |
| R-R-R | 2007-2021 | -9.5 | -11.6 | -7.3 | < 0.0001 |
| R-S-R | 2007-2021 | -2.7 | -6 | 0.6 | 0.1110 |
| S-S-R | 2007-2021 | -0.6 | -2.9 | 1.6 | 0.5785 |
| S-S-S | 2007-2021 | -3.8 | -6 | -1.5 | 0.0012 |
| S-R-R | 2007-2021 | -2 | -5.4 | 1.6 | 0.2751 |
| R-R-S | 2007-2021 | -3.2 | -6.1 | -0.2 | 0.0379 |

Table S5. Multinomial logistic regression results for *Staphylococcus aureus* patchwork dataset. Estimates are presented as the percentage change in the odds of each resistance phenotype for every additional treatment day (per 100 patient-days) of exposure to the specified antimicrobial class in the preceding 14 days, compared with isolates fully susceptible to all key classes (S-S-S phenotype). Antibiotic classes used for phenotypes: fluoroquinolones, anti-staphylococcal beta-lactams, macrolides. This table corresponds to Figure 3A in the main text.

| **Antibiotic exposure** | **Outcome phenotype** | **Estimate** | **80% confidence interval** | | **95% confidence interval** | | **p-value** |
| --- | --- | --- | --- | --- | --- | --- | --- |
|  |  |  | **Lower bound** | **Upper bound** | **Lower bound** | **Upper bound** |  |
| **Anti-staphylococcal Beta-lactams** | R-R-R | 0.3 | -1.4 | 1.9 | -2.2 | 2.8 | 0.8394 |
|  | S-S-R | -1.8 | -4.1 | 0.6 | -5.3 | 1.9 | 0.3430 |
|  | S-R-R | -2.1 | -4.9 | 0.9 | -6.4 | 2.4 | 0.3621 |
|  | R-S-R | -0.7 | -3.8 | 2.5 | -5.3 | 4.2 | 0.7800 |
|  | R-R-S | -4.6 | -8.2 | -0.8 | -10.1 | 1.3 | 0.1257 |
| **Macrolides** | R-R-R | 1.4 | -0.3 | 3.1 | -1.2 | 4 | 0.2927 |
|  | S-S-R | 0.7 | -1.7 | 3.2 | -3 | 4.6 | 0.7029 |
|  | **S-R-R** | **4.5** | **1.8** | **7.3** | **0.4** | **8.8** | **0.0302** |
|  | R-S-R | -2.7 | -6 | 0.6 | -7.6 | 2.5 | 0.2982 |
|  | R-R-S | -3.6 | -7.5 | 0.5 | -9.5 | 2.7 | 0.2553 |
| **Fluoroquinolones** | **R-R-R** | **1.7** | **0.8** | **2.5** | **0.4** | **3** | **0.0127** |
|  | S-S-R | 1.2 | -0.1 | 2.5 | -0.8 | 3.2 | 0.2405 |
|  | S-R-R | -0.7 | -2.2 | 0.8 | -2.9 | 1.6 | 0.5443 |
|  | **R-S-R** | **3** | **1.4** | **4.7** | **0.6** | **5.6** | **0.0155** |
|  | **R-R-S** | **4.6** | **2.6** | **6.6** | **1.5** | **7.7** | **0.0033** |

Table S6. Multinomial logistic regression results for *Staphylococcus aureus* patchwork dataset for community prevalence covariates. Estimates are presented as the percentage change in the odds of each resistance phenotype for each increase in community prevalence of the specified antimicrobial class in the preceding 14 days, compared with isolates fully susceptible to all key classes (S-S-S phenotype). Community prevalence was measured by the incidence rate of phenotypes collected within 3 days of admission during the previous 90 days (referred to as importation phenotype). Antibiotic classes used for phenotypes: fluoroquinolones, anti-staphylococcal beta-lactams, macrolides. This table corresponds to Figure S1.

| **Outcome phenotype** | **Importation phenotype** | **Estimate** | **80% confidence interval** | | **95% confidence interval** | | **p-value** |
| --- | --- | --- | --- | --- | --- | --- | --- |
|  |  |  | **Lower bound** | **Upper bound** | **Lower bound** | **Upper bound** |  |
| R-R-R | **R-R-R** | **1.1** | **0.8** | **1.5** | **0.6** | **1.6** | **< 0.0001** |
|  | S-S-S | 0.2 | -0.2 | 0.5 | -0.4 | 0.7 | 0.5651 |
|  | S-S-R | 0.6 | 0.2 | 1.1 | 0 | 1.3 | 0.0687 |
|  | S-R-R | 0.4 | -0.1 | 0.8 | -0.3 | 1.1 | 0.2900 |
|  | **R-S-R** | **1.3** | **0.7** | **1.9** | **0.3** | **2.2** | **0.0072** |
|  | **R-R-S** | **1.3** | **0.5** | **2.1** | **0.1** | **2.5** | **0.0290** |
| S-S-R | R-R-R | -0.2 | -0.7 | 0.2 | -1 | 0.5 | 0.4961 |
|  | S-S-S | -0.6 | -1.1 | -0.1 | -1.4 | 0.2 | 0.1304 |
|  | S-S-R | -0.1 | -0.8 | 0.5 | -1.1 | 0.9 | 0.7838 |
|  | S-R-R | 0.1 | -0.5 | 0.8 | -0.9 | 1.1 | 0.8068 |
|  | R-S-R | 1.1 | 0.2 | 2 | -0.2 | 2.4 | 0.1095 |
|  | R-R-S | 1.3 | 0.2 | 2.4 | -0.4 | 3 | 0.1452 |
| S-R-R | R-R-R | -0.3 | -0.9 | 0.3 | -1.2 | 0.6 | 0.5175 |
|  | S-S-S | -0.3 | -0.9 | 0.4 | -1.2 | 0.7 | 0.5722 |
|  | S-S-R | 0.5 | -0.3 | 1.3 | -0.7 | 1.7 | 0.4141 |
|  | **S-R-R** | **2.2** | **1.5** | **3** | **1.1** | **3.4** | **0.0001** |
|  | R-S-R | 1.1 | 0.1 | 2.2 | -0.5 | 2.8 | 0.1686 |
|  | **R-R-S** | 0.9 | -0.5 | 2.3 | -1.2 | 3.1 | 0.3971 |
| R-S-R | R-R-R | 1.7 | 1 | 2.3 | 0.7 | 2.7 | 0.0010 |
|  | S-S-S | 0.8 | 0.1 | 1.5 | -0.3 | 1.9 | 0.1525 |
|  | S-S-R | 1.1 | 0.2 | 2 | -0.3 | 2.5 | 0.1296 |
|  | S-R-R | 1.3 | 0.4 | 2.3 | 0 | 2.8 | 0.0577 |
|  | **R-S-R** | **3** | **1.9** | **4.2** | **1.2** | **4.8** | **0.0008** |
|  | **R-R-S** | **2.4** | **0.8** | **3.9** | **0** | **4.7** | **0.0469** |
| R-R-S | **R-R-R** | **2.6** | **1.8** | **3.4** | **1.3** | **3.8** | **< 0.0001** |
|  | S-S-S | 1.3 | 0.5 | 2.2 | 0 | 2.7 | 0.0501 |
|  | **S-S-R** | **2** | **0.9** | **3.1** | **0.3** | **3.7** | **0.0184** |
|  | S-R-R | 0.1 | -1 | 1.2 | -1.6 | 1.8 | 0.9171 |
|  | **R-S-R** | **2.7** | **1.3** | **4.2** | **0.6** | **4.9** | **0.0137** |
|  | **R-R-S** | **6.6** | **4.9** | **8.2** | **4.1** | **9.1** | **< 0.0001** |

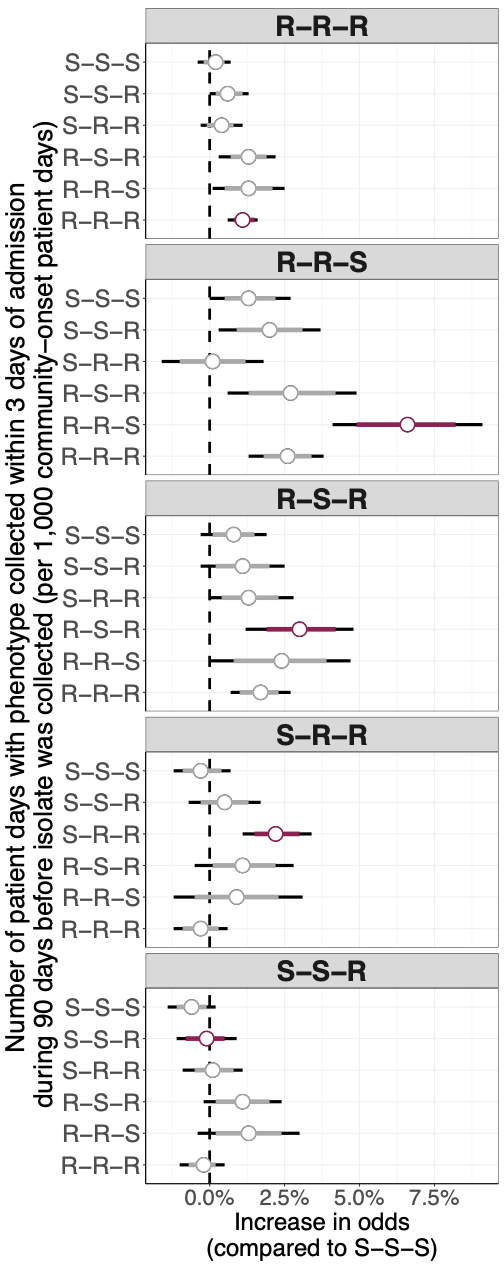

Figure S3. Multinomial logistic regression results for community prevalence covariates for *Staphylococcus aureus*. Points represent coefficient estimates and line intervals represent confidence intervals (80% in red and 95% in black). Estimates are presented as the percentage change in the odds of each resistance phenotype for each increase in community prevalence of the specified antimicrobial class in the preceding 14 days, compared with isolates fully susceptible to all key classes (S-S-S phenotype). Community prevalence was measured by the incidence rate of phenotypes collected within 3 days of admission during the previous 90 days (referred to as importation phenotype). Antibiotic classes used for phenotypes: fluoroquinolones, anti-staphylococcal beta-lactams, macrolides.

#### S3.2  *Escherichia coli*

##### S3.2.1 Main results

Table S4. Most common phenotypes for *Escherichia coli* clinical 30-day incident isolates in the patchwork dataset, from Feb 1, 2007 to Dec 31, 2021. The phenotype in bold was used as the reference category (S-S-S) in the multinomial logistic regression analysis.

|  | **Fluoroquinolones** | **3^rd^ and 4^th^ generation cephalosporins** | **Beta-lactam/**  **Beta-lactamase inhibitors** | **Count** | **Percentage (%)** | **Cumulative percentage (%)** | **Abbreviation** |
| --- | --- | --- | --- | --- | --- | --- | --- |
| **1** | **S** | **S** | **S** | **13334** | **42.8** | **42.8** | **S-S-S** |
| 2 | R | S | R | 5272 | 16.9 | 59.7 | R-S-R |
| 3 | S | S | R | 5139 | 16.5 | 76.2 | S-S-R |
| 4 | R | S | S | 3034 | 9.7 | 85.9 | R-S-S |
| 5 | R | R | R | 3007 | 9.6 | 95.5 | R-R-R |
| 6 | S | R | R | 709 | 2.3 | 97.8 | S-R-R |
| 7 | R | R | S | 380 | 1.2 | 99.0 | R-R-S |

Table S5. Time trend estimates for *E coli* phenotype incidence, from Feb 1, 2007 to Dec 31, 2021. Time trend estimates represent average annual percentage change (AAPC) and were obtained using generalized estimating equations. This table corresponds to Figure 2B in the main text.

| **Phenotype** | **Time period** | **Time trend estimate** | **Lower 95% confidence bound** | **Upper 95% confidence bound** | **p-value** |
| --- | --- | --- | --- | --- | --- |
| **R-S-R** | **2007-2021** | **-5.6** | **-7.6** | **-3.7** | **< 0.0001** |
| **R-S-S** | **2007-2021** | **-8** | **-11.3** | **-4.6** | **< 0.0001** |
| S-S-R | 2007-2021 | -1.8 | -4.2 | 0.6 | = 0.1411 |
| **S-S-S** | **2007-2021** | **-5.1** | **-6.8** | **-3.5** | **< 0.0001** |
| R-R-R | 2007-2021 | 2.2 | -0.6 | 5.1 | = 0.1272 |
| S-R-R | 2007-2021 | 0.8 | -2.3 | 3.9 | = 0.6281 |
| **R-R-S** | **2007-2021** | **4.9** | **0.6** | **9.4** | **= 0.0264** |

Table S6. Multinomial logistic regression results for *Escherichia coli* patchwork dataset. Estimates are presented as the percentage change in the odds of each resistance phenotype for every additional treatment day (per 100 patient-days) of exposure to the specified antimicrobial class in the preceding 14 days, compared with isolates fully susceptible to all key classes (S-S-S phenotype). Antibiotic classes used for phenotypes: fluoroquinolones, 3^rd^ and 4^th^ generation cephalosporins, and beta-lactam/beta-lactamase inhibitors. This table corresponds to Figure 3B in the main text.

| **Antibiotic exposure** | **Outcome**  **phenotype** | **Estimate** | **80% confidence interval** | | **95% confidence interval** | | **p-value** |
| --- | --- | --- | --- | --- | --- | --- | --- |
|  |  |  | **Lower bound** | **Upper bound** | **Lower bound** | **Upper bound** |  |
| **Fluoroquinolones** | **R-S-R** | **3.6** | **2.6** | **4.6** | **2.1** | **5.2** | **< 0.0001** |
|  | S-S-R | -0.4 | -1.5 | 0.6 | -2 | 1.2 | 0.5853 |
|  | **R-S-S** | **3.8** | **2.7** | **4.8** | **2.2** | **5.4** | **< 0.0001** |
|  | **R-R-R** | **2.9** | **1.8** | **4.1** | **1.1** | **4.7** | **0.0012** |
|  | S-R-R | -1 | -3.1 | 1.1 | -4.2 | 2.2 | 0.5247 |
|  | R-R-S | 4.8 | 2 | 7.8 | 0.5 | 9.4 | 0.0298 |
| **3^rd^ and 4^th^ generation cephalosporins** | R-S-R | 0.2 | -0.8 | 1.2 | -1.3 | 1.7 | 0.8019 |
|  | S-S-R | 1.3 | 0.4 | 2.3 | -0.1 | 2.8 | 0.0787 |
|  | R-S-S | -0.2 | -1.2 | 0.8 | -1.7 | 1.4 | 0.8082 |
|  | **R-R-R** | **2** | **1** | **3** | **0.5** | **3.5** | **0.0102** |
|  | **S-R-R** | **4.8** | **3** | **6.6** | **2.1** | **7.5** | **0.0004** |
|  | R-R-S | -1.5 | -3.8 | 0.9 | -5 | 2.1 | 0.4110 |
| **Beta-lactam/ Beta-lactamase inhibitors** | **R-S-R** | **1.3** | **0.5** | **2.2** | **0** | **2.6** | **0.0465** |
|  | **S-S-R** | **2.2** | **1.4** | **3.1** | **0.9** | **3.5** | **0.0006** |
|  | R-S-S | 0.4 | -0.4 | 1.2 | -0.9 | 1.7 | 0.5580 |
|  | R-R-R | 0 | -0.9 | 0.9 | -1.4 | 1.3 | 0.9492 |
|  | S-R-R | **3.1** | **1.5** | **4.8** | **0.7** | **5.7** | **0.0130** |
|  | R-R-S | -0.8 | -2.8 | 1.3 | -3.8 | 2.4 | 0.6268 |

Table S7. Multinomial logistic regression results for *Escherichia coli* patchwork dataset for community prevalence covariates. Estimates are presented as the percentage change in the odds of each resistance phenotype for each increase in community prevalence of the specified antimicrobial class in the preceding 14 days, compared with isolates fully susceptible to all key classes (S-S-S phenotype). Community prevalence was measured by the incidence rate of phenotypes collected within 3 days of admission during the previous 90 days (referred to as importation phenotype). Antibiotic classes used for phenotypes: fluoroquinolones, 3^rd^ and 4^th^ generation cephalosporins, and beta-lactam/beta-lactamase inhibitors. This table corresponds to Figure S2.

| **Outcome phenotype** | **Importation phenotype** | **Estimate** | **80% confidence interval** | | **95% confidence interval** | | **p-value** |
| --- | --- | --- | --- | --- | --- | --- | --- |
|  |  |  | **Lower bound** | **Upper bound** | **Lower bound** | **Upper bound** |  |
| R-S-R | **S-S-S** | **-0.6** | **-0.9** | **-0.3** | **-1.1** | **-0.1** | **0.0156** |
|  | **R-S-R** | **1.7** | **1.3** | **2.2** | **1** | **2.4** | **< 0.0001** |
|  | **S-S-R** | **1.4** | **0.9** | **1.9** | **0.7** | **2.1** | 0.0002 |
|  | R-S-S | -1.9 | -2.3 | -1.5 | -2.5 | -1.2 | **< 0.0001** |
|  | R-R-R | 0.2 | -0.4 | 0.7 | -0.7 | 1 | 0.7009 |
|  | S-R-R | 0.8 | -0.4 | 2 | -1.1 | 2.6 | 0.4160 |
|  | **R-R-S** | **-3.7** | **-4.9** | **-2.4** | **-5.6** | **-1.8** | **0.0002** |
| S-S-R | S-S-S | -0.5 | -0.8 | -0.2 | -0.9 | 0 | 0.0545 |
|  | **R-S-R** | **1.5** | **1** | **2** | **0.8** | **2.2** | **< 0.0001** |
|  | **S-S-R** | **1.5** | **1** | **1.9** | **0.8** | **2.2** | **< 0.0001** |
|  | R-S-S | -1.9 | -2.4 | -1.5 | -2.6 | -1.3 | **< 0.0001** |
|  | R-R-R | 0.7 | 0.2 | 1.3 | -0.1 | 1.5 | 0.0750 |
|  | S-R-R | -1.4 | -2.5 | -0.2 | -3.2 | 0.5 | 0.1463 |
|  | **R-R-S** | **-3.6** | **-4.8** | **-2.4** | **-5.4** | **-1.8** | 0.0001 |
| R-S-S | **S-S-S** | **0.7** | **0.4** | **1.1** | **0.2** | **1.2** | **0.0044** |
|  | R-S-R | 0.3 | -0.2 | 0.8 | -0.5 | 1.1 | 0.4638 |
|  | S-S-R | -0.4 | -0.9 | 0.2 | -1.2 | 0.5 | 0.3992 |
|  | **R-S-S** | **2.3** | **1.9** | **2.7** | **1.7** | **2.9** | **< 0.0001** |
|  | R-R-R | -0.3 | -0.9 | 0.3 | -1.2 | 0.6 | 0.5501 |
|  | S-R-R | 1.7 | 0.4 | 2.9 | -0.2 | 3.6 | 0.0822 |
|  | R-R-S | 1.1 | 0.2 | 2.1 | -0.3 | 2.6 | 0.1178 |
| R-R-R | S-S-S | -0.1 | -0.5 | 0.2 | -0.7 | 0.4 | 0.5859 |
|  | **R-S-R** | **1.1** | **0.6** | **1.6** | **0.3** | **1.9** | 0.0058 |
|  | S-S-R | 0.8 | 0.3 | 1.4 | 0 | 1.6 | 0.0435 |
|  | R-S-S | -1 | -1.5 | -0.5 | -1.7 | -0.3 | 0.0071 |
|  | **R-R-R** | **2.2** | **1.6** | **2.7** | **1.4** | **3** | **< 0.0001** |
|  | S-R-R | 1.3 | 0.1 | 2.5 | -0.5 | 3.1 | 0.1530 |
|  | **R-R-S** | **-4** | **-5.3** | **-2.8** | **-5.9** | **-2.1** | **< 0.0001** |
| S-R-R | S-S-S | 0 | -0.7 | 0.6 | -1 | 1 | 0.9625 |
|  | R-S-R | 1.5 | 0.5 | 2.4 | 0 | 3 | 0.0540 |
|  | S-S-R | 0.2 | -0.8 | 1.2 | -1.3 | 1.7 | 0.7888 |
|  | R-S-S | -1 | -1.9 | -0.1 | -2.3 | 0.3 | 0.1428 |
|  | R-R-R | 1.4 | 0.4 | 2.4 | -0.2 | 2.9 | 0.0810 |
|  | **S-R-R** | **5.5** | **3.8** | **7.3** | **2.9** | **8.2** | **< 0.0001** |
|  | R-R-S | -1.6 | -3.8 | 0.7 | -5 | 1.9 | 0.3616 |
| R-R-S | S-S-S | 0.4 | -0.4 | 1.2 | -0.9 | 1.6 | 0.5528 |
|  | R-S-R | -2.3 | -3.7 | -0.8 | -4.5 | 0 | 0.0517 |
|  | S-S-R | -0.3 | -1.6 | 1.1 | -2.3 | 1.8 | 0.7922 |
|  | **R-S-S** | **1.6** | **0.6** | **2.6** | **0.1** | **3.1** | **0.0345** |
|  | R-R-R | -4.5 | -6 | -3 | -6.8 | -2.2 | 0.0002 |
|  | S-R-R | -1.4 | -4.5 | 1.7 | -6.1 | 3.5 | 0.5593 |
|  | **R-R-S** | **6.5** | **4.9** | **8.1** | **4.1** | **9** | **< 0.0001** |

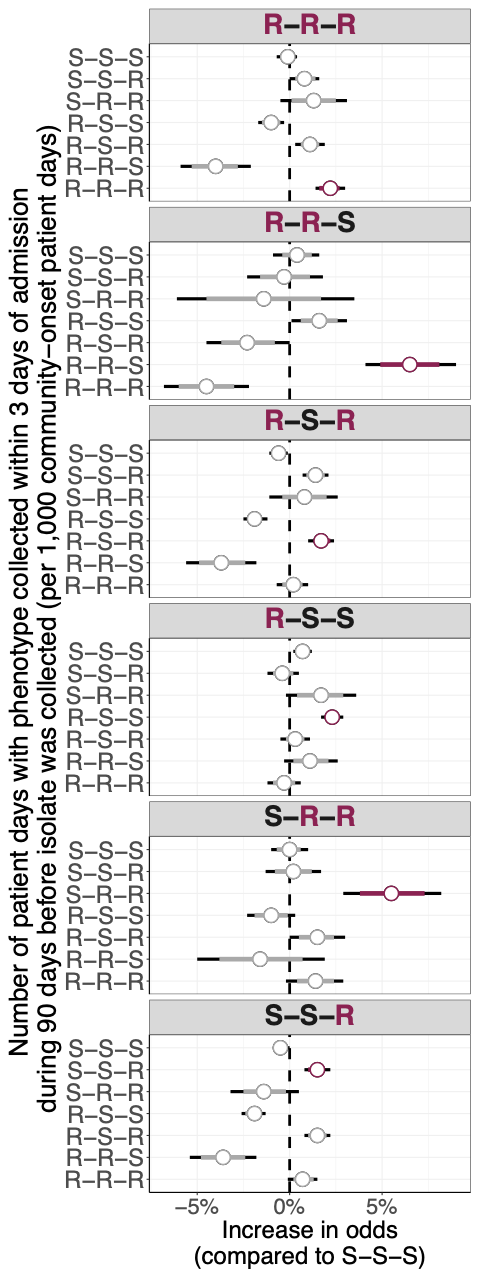

Figure S4. Multinomial logistic regression results for community prevalence covariates for *Escherichia coli*. Points represent coefficient estimates and line intervals represent confidence intervals (80% in red and 95% in black). Estimates are presented as the percentage change in the odds of each resistance phenotype for each increase in community prevalence of the specified antimicrobial class in the preceding 14 days, compared with isolates fully susceptible to all key classes (S-S-S phenotype). Community prevalence was measured by the incidence rate of phenotypes collected within 3 days of admission during the previous 90 days (referred to as importation phenotype). Antibiotic classes used for phenotypes: fluoroquinolones, 3^rd^/4^th^ generation cephalosporins, beta-lactam/beta-lactamase inhibitors.

##### S3.2.2 Sensitivity analysis with 3^rd^ and 4^th^ generation cephalosporins as separate categories

Table S8. Most common four-drug phenotypes for *Escherichia coli* clinical 30-day incident isolates in the patchwork dataset, from Feb 1, 2007 to Dec 31, 2021. The phenotype in bold was used as the reference category (S-S-S-S) in the multinomial logistic regression analysis.

|  | **Fluoroquinolones** | **3^rd^ generation cephalosporins** | **4^th^ generation cephalosporins** | **Beta-lactam/**  **Beta-lactamase inhibitors** | **Count** | **Percentage (%)** | **Cumulative percentage (%)** | **Abbreviation** |
| --- | --- | --- | --- | --- | --- | --- | --- | --- |
| **1** | **S** | **S** | **S** | **S** | **10085** | **41.6** | **41.6** | **S-S-S-S** |
| 2 | S | S | S | R | 4172 | 17.2 | 58.9 | S-S-S-R |
| 3 | R | S | S | R | 4159 | 17.2 | 76.0 | R-S-S-R |
| 4 | R | S | S | S | 2160 | 8.9 | 84.9 | R-S-S-S |
| 5 | R | R | R | R | 2160 | 8.9 | 93.9 | R-R-R-R |
| 6 | R | R | S | R | 328 | 1.4 | 95.2 | R-R-S-R |

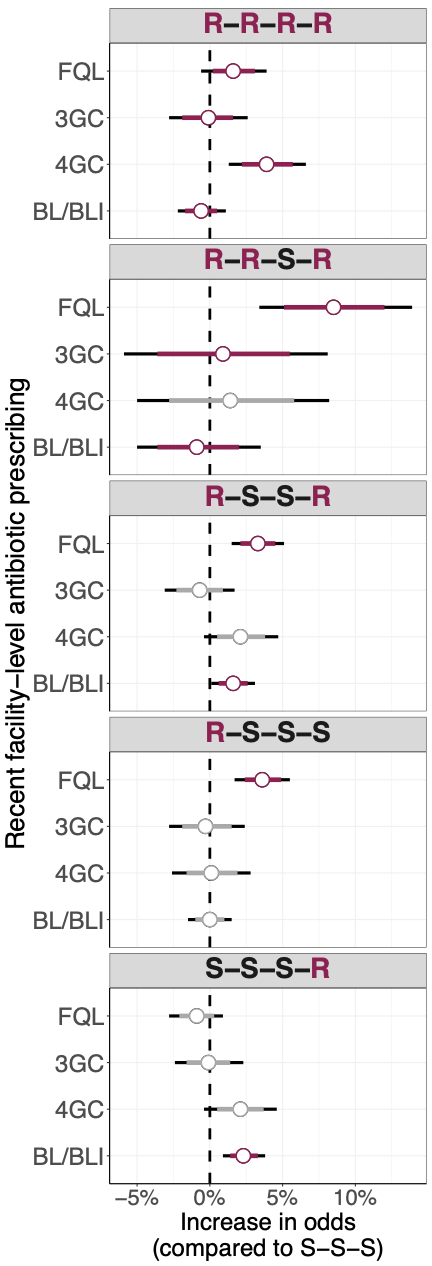

Figure S5. Effect of recent antimicrobial prescribing on four-drug resistance patterns for *Escherichia coli*. Patchwork data was used for the analysis and the approach is described in the appendix p.28. Multilevel multinomial logistic regression coefficients are shown as the percentage change in the odds of each resistance phenotype for every additional treatment day (per 100 patient-days) of exposure to the specified antimicrobial class in the preceding 14 days (per 1,000 patient-days), compared with isolates fully susceptible to all key classes (S-S-S). Points denote medians; thick bars, 80 % confidence intervals; thin bars, 95 % confidence intervals. Coefficients highlighted in dark red correspond to antimicrobial-pathogen combinations for which an effect of prescribing on resistance was hypothesised. Antibiotic classes: FQL = fluoroquinolones, 3GC = 3^rd^ generation cephalosporins, 4GC = 4th generation cephalosporins, BL/BLI = beta-lactam/beta-lactamase inhibitors.

#### S3.3  *Klebsiella pneumoniae*

##### S3.3.1  Main results

Table S9. Most common phenotypes for *Klebsiella pneumoniae* clinical 30-day incident isolates in the patchwork dataset, from Feb 1, 2007 to Dec 31, 2021. The phenotype in bold was used as the reference category (S-S-S) in the multinomial logistic regression analysis.

|  | **Fluoroquinolones** | **3^rd^ and 4^th^ generation cephalosporins** | **Beta-lactam/**  **Beta-lactamase inhibitors** | **Count** | | **Percentage (%)** | **Cumulative percentage (%)** | **Abbreviation** |
| --- | --- | --- | --- | --- | --- | --- | --- | --- |
| **1** | S | S | S | 17634 | 66.5 | | 66.5 | S-S-S |
| 2 | S | S | R | 3329 | 12.6 | | 79.0 | S-S-R |
| 3 | R | R | R | 3255 | 12.3 | | 91.3 | R-R-R |
| 4 | S | R | R | 1050 | 4.0 | | 95.3 | S-R-R |
| 5 | R | S | R | 473 | 1.8 | | 97.0 | R-S-R |

Table S10. Time trend estimates for *Klebsiella pneumoniae* phenotype incidence, from Feb 1, 2007 to Dec 31, 2021. Time trend estimates represent average annual percentage change (AAPC) and were obtained using generalized estimating equations. This table corresponds to Figure 2B in the main text.

| **Phenotype** | **Time period** | **Time trend estimate** | **Lower 95% confidence bound** | **Upper 95% confidence bound** | **p-value** |
| --- | --- | --- | --- | --- | --- |
| **S-S-S** | **2007-2021** | **-5.5** | **-7.1** | **-4** | **< 0.0001** |
| S-S-R | 2007-2021 | 0 | -2.8 | 2.8 | 0.9809 |
| S-R-R | 2007-2021 | 1.2 | -1.5 | 4 | 0.3903 |
| **R-R-R** | **2007-2021** | **-5.5** | **-10** | **-0.6** | **0.0275** |
| **R-S-R** | **2007-2021** | **-6** | **-8.5** | **-3.5** | **< 0.0001** |
| **S-S-S** | **2007-2021** | **-5.5** | **-7.1** | **-4** | **< 0.0001** |
| **S-S-R** | **2007-2021** | **0** | **-2.8** | **2.8** | **0.9809** |

Table S11. Multinomial logistic regression results for *Klebsiella pneumoniae* patchwork dataset. Estimates are presented as the percentage change in the odds of each resistance phenotype for every additional treatment day (per 100 patient-days) of exposure to the specified antimicrobial class in the preceding 14 days, compared with isolates fully susceptible to all key classes (S-S-S phenotype). Antibiotic classes used for phenotypes: fluoroquinolones, 3^rd^ generation cephalosporins, and beta-lactam/beta-lactamase inhibitors. This table corresponds to Figure 3C in the main text.

| **Term** | **Outcome phenotype** | **Estimate** | **80% confidence interval** | | **95% confidence interval** | | **p-value** |
| --- | --- | --- | --- | --- | --- | --- | --- |
|  |  |  | **Lower bound** | **Upper bound** | **Lower bound** | **Upper bound** |  |
| **Fluoroquinolones** | S-S-R | 0.5 | -0.9 | 1.9 | -1.6 | 2.6 | 0.5353 |
|  | **R-R-R** | **2.3** | **1.1** | **3.6** | **0.4** | **4.3** | **0.0094** |
|  | S-R-R | -1.1 | -2.9 | 0.7 | -3.8 | 1.6 | 0.5122 |
|  | **R-S-R** | **6.3** | **3.3** | **9.3** | **1.8** | **11** | **0.0034** |
| **3^rd^ and 4^th^ generation cephalosporins** | S-S-R | 0.4 | -1.3 | 2.1 | -2.2 | 3.1 | 0.3911 |
|  | **R-R-R** | **0.4** | **-1.1** | **2.1** | **-2** | **2.9** | **0.0002** |
|  | **S-R-R** | **3** | **0.9** | **5.1** | **-0.1** | **6.2** | **0.0002** |
|  | **R-S-R** | **-7.5** | **-11.8** | **-3.1** | **-14** | **-0.6** | **0.0003** |
| **Beta-lactam/ Beta-lactamase inhibitors** | **S-S-R** | **2.5** | **1.4** | **3.6** | **0.9** | **4.1** | **0.0027** |
|  | R-R-R | -1.1 | -2 | -0.2 | -2.5 | 0.3 | 0.2565 |
|  | **S-R-R** | **2.2** | **0.9** | **3.5** | **0.2** | **4.2** | **0.0033** |
|  | R-S-R | 2.5 | -0.1 | 5 | -1.4 | 6.4 | 0.5001 |

Table S12. Multinomial logistic regression results for *Klebsiella pneumoniae* patchwork dataset for community prevalence covariates. Estimates are presented as the percentage change in the odds of each resistance phenotype for each increase in community prevalence of the specified antimicrobial class in the preceding 14 days, compared with isolates fully susceptible to all key classes (S-S-S phenotype). Community prevalence was measured by the incidence rate of phenotypes collected within 3 days of admission during the previous 90 days (referred to as importation phenotype). Antibiotic classes used for phenotypes: fluoroquinolones, 3^rd^ and 4^th^ generation cephalosporins, and beta-lactam/beta-lactamase inhibitors. This table corresponds to Figure S3.

| **Outcome phenotype** | **Importation phenotype** | **Estimate** | **80% confidence interval** | | **95% confidence interval** | | **p-value** |
| --- | --- | --- | --- | --- | --- | --- | --- |
|  |  |  | **Lower bound** | **Upper bound** | **Lower bound** | **Upper bound** |  |
| S-S-R | **S-S-S** | **-0.5** | **-0.9** | **-0.2** | **-1.1** | **0** | 0.0451 |
|  | **S-S-R** | **-0.4** | **-0.7** | **-0.1** | **-0.9** | **0** | 0.0694 |
|  | R-R-R | 0.3 | -0.2 | 0.7 | -0.4 | 1 | 0.4207 |
|  | S-R-R | -0.7 | -1.6 | 0.1 | -2 | 0.5 | 0.2490 |
|  | R-S-R | 1.1 | 0.4 | 1.8 | 0.1 | 2.1 | 0.0345 |
| R-R-R | S-S-S | -0.5 | -1.1 | 0.1 | -1.5 | 0.5 | 0.2985 |
|  | S-S-R | 1.1 | 0.3 | 2 | -0.2 | 2.5 | 0.0918 |
|  | R-R-R | 0.1 | -1.6 | 1.8 | -2.5 | 2.7 | 0.9505 |
|  | S-R-R | 0 | -0.5 | 0.6 | -0.8 | 0.9 | 0.9500 |
|  | **R-S-R** | **1.5** | **1** | **1.9** | **0.8** | **2.1** | < 0.0001 |
| S-R-R | S-S-S | 1 | 0.4 | 1.7 | 0 | 2.1 | 0.0417 |
|  | S-S-R | 0.4 | -0.8 | 1.6 | -1.4 | 2.3 | 0.6606 |
|  | R-R-R | -0.2 | -1 | 0.7 | -1.5 | 1.2 | 0.8139 |
|  | S-R-R | -0.1 | -0.9 | 0.6 | -1.2 | 1 | 0.8105 |
|  | **R-S-R** | **4.7** | **3.9** | **5.5** | **3.5** | **5.9** | < 0.0001 |
| R-S-R | S-S-S | -1.3 | -3.5 | 1.1 | -4.7 | 2.3 | 0.4857 |
|  | S-S-R | 0.3 | -1.2 | 1.8 | -2 | 2.7 | 0.8073 |
|  | R-R-R | 2.7 | 1.6 | 3.8 | 1 | 4.4 | 0.0021 |
|  | S-R-R | -0.3 | -2.3 | 1.7 | -3.4 | 2.8 | 0.8311 |
|  | R-S-R | 1.2 | -1.9 | 4.4 | -3.5 | 6.1 | 0.6320 |

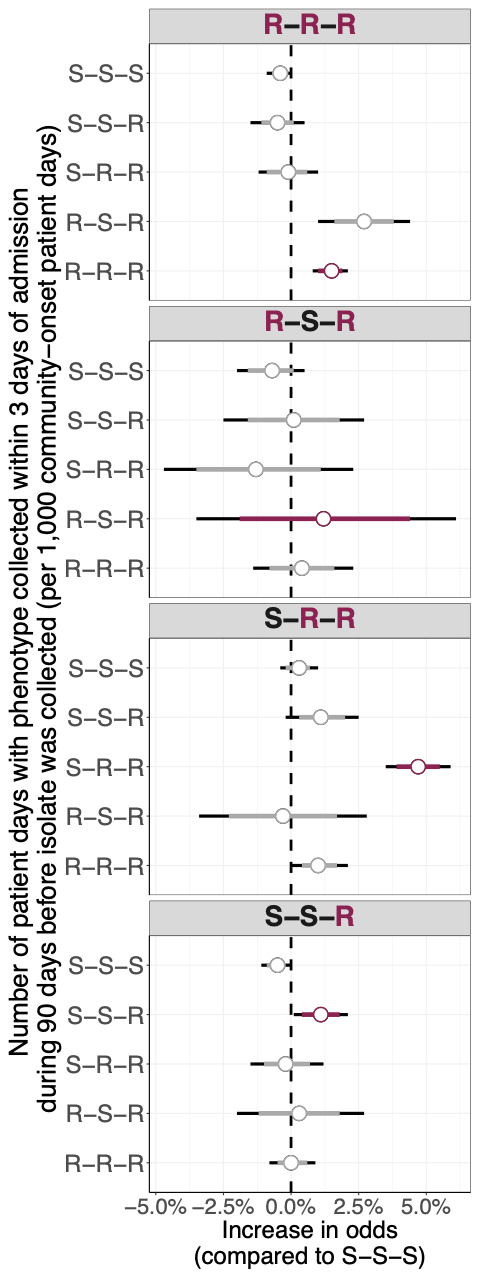

Figure S3. Multinomial logistic regression results for community prevalence covariates for *Klebsiella pneumoniae*. Points represent coefficient estimates and line intervals represent confidence intervals (80% in red and 95% in black). Estimates are presented as the percentage change in the odds of each resistance phenotype for each increase in community prevalence of the specified antimicrobial class in the preceding 14 days, compared with isolates fully susceptible to all key classes (S-S-S phenotype). Community prevalence was measured by the incidence rate of phenotypes collected within 3 days of admission during the previous 90 days (referred to as importation phenotype). Antibiotic classes used for phenotypes: fluoroquinolones, 3^rd^ and 4^th^ generation cephalosporins, beta-lactam/beta-lactamase inhibitors.

##### S3.3.2  Sensitivity analysis with 3^rd^ and 4^th^ generation cephalosporins as separate categories

Table S13. Most common four-drug phenotypes for *Klebsiella pneumoniae* clinical 30-day incident isolates in the patchwork dataset, from Feb 1, 2007 to Dec 31, 2021. The phenotype in bold was used as the reference category (S-S-S-S) in the multinomial logistic regression analysis.

|  | **Fluoroquinolones** | **3^rd^ generation cephalosporins** | **4^th^ generation cephalosporins** | **Beta-lactam/**  **Beta-lactamase inhibitors** | **Count** | **Percentage (%)** | **Cumulative percentage (%)** | **Abbreviation** |
| --- | --- | --- | --- | --- | --- | --- | --- | --- |
| **1** | **S** | **S** | **S** | **S** | **13,668** | **65.4** | **65.4** | **S-S-S-S** |
| 2 | S | S | S | R | 2,706 | 13.0 | 78.4 | S-S-S-R |
| 3 | R | R | R | R | 2,550 | 12.2 | 90.6 | R-R-R-R |
| 4 | S | R | R | R | 608 | 2.9 | 93.5 | S-R-R-R |
| 5 | R | S | S | R | 345 | 1.7 | 95.1 | R-S-S-R |

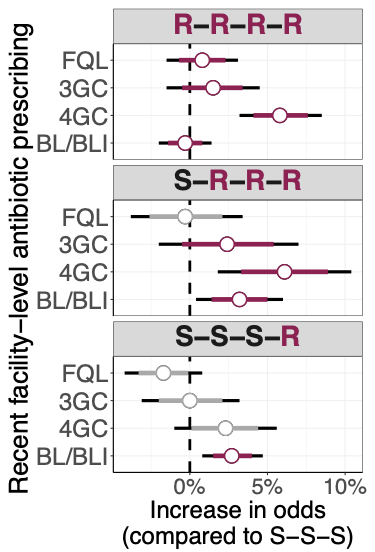

Figure S6. Effect of recent antimicrobial prescribing on four-drug resistance patterns for *Klebsiella pneumoniae*. Patchwork data was used for the analysis, and the approach is described in the appendix p.28. Multilevel multinomial logistic regression coefficients are shown as the percentage change in the odds of each resistance phenotype for every additional treatment day (per 100 patient-days) of exposure to the specified antimicrobial class in the preceding 14 days (per 1,000 patient-days), compared with isolates fully susceptible to all key classes (S-S-S). Points denote medians; thick bars, 80 % confidence intervals; thin bars, 95 % confidence intervals. Coefficients highlighted in dark red correspond to antimicrobial-pathogen combinations for which an effect of prescribing on resistance was hypothesised. Antibiotic classes: FQL = fluoroquinolones, 3GC = 3^rd^ generation cephalosporins, 4GC = 4th generation cephalosporins, BL/BLI = beta-lactam/beta-lactamase inhibitors.

#### S3.4  *Pseudomonas aeruginosa*

Table S15. Most common phenotypes for *Pseudomonas aeruginosa* clinical 30-day incident isolates in the patchwork dataset, from Feb 1, 2007 to Dec 31, 2021. The phenotype in bold was used as the reference category (S-S-S) in the multinomial logistic regression analysis.

|  | **Fluoroquinolones** | **Beta-lactam/**  **Beta-lactamase inhibitors** | **Carbapenems** | **Count** | **Percentage (%)** | **Cumulative percentage (%)** | **Abbreviation** |
| --- | --- | --- | --- | --- | --- | --- | --- |
| **1** | **S** | **S** | **S** | **10928** | **47.4** | **47.4** | **S-S-S** |
| 2 | S | R | S | 3080 | 13.3 | 60.7 | S-R-S |
| 3 | R | R | R | 2201 | 9.5 | 70.2 | R-R-R |
| 4 | R | S | S | 2000 | 8.7 | 78.9 | R-S-S |
| 5 | R | R | S | 1633 | 7.1 | 86 | R-R-S |
| 6 | R | S | R | 1152 | 5 | 91 | R-S-R |
| 7 | S | S | R | 920 | 4 | 95 | S-S-R |

Table S16. Time trend estimates for *Pseudomonas aeruginosa* phenotype incidence, from Feb 1, 2007 to Dec 31, 2021. Time trend estimates represent average annual percentage change (AAPC) and were obtained using generalized estimating equations. This table corresponds to Figure 2B in the main text.

| **Phenotype** | **Time period** | **Time trend estimate** | **95% confidence interval** | | **p-value** |
| --- | --- | --- | --- | --- | --- |
|  |  |  | **Lower bound** | **Upper bound** |  |
| R-S-S | 2007-2021 | -10 | -13.2 | -6.7 | < 0.0001 |
| S-R-S | 2007-2021 | -8.7 | -13.2 | -4.1 | = 4e-04 |
| S-S-S | 2007-2021 | -2.6 | -4.9 | -0.2 | = 0.031 |
| R-R-R | 2007-2021 | -8.7 | -12.8 | -4.5 | < 0.0001 |
| R-R-S | 2007-2021 | -7.8 | -12.5 | -2.9 | = 0.0024 |
| R-S-R | 2007-2021 | -6.4 | -10.3 | -2.4 | = 0.0023 |
| S-S-R | 2007-2021 | -1.7 | -4.5 | 1.1 | = 0.2257 |

Table S17. Multinomial logistic regression results for *Pseudomonas aeruginosa* patchwork dataset. Estimates are presented as the percentage change in the odds of each resistance phenotype for every additional treatment day (per 100 patient-days) of exposure to the specified antimicrobial class in the preceding 14 days, compared with isolates fully susceptible to all key classes (S-S-S phenotype). Antibiotic classes used for phenotypes: fluoroquinolones, beta-lactam/beta-lactamase inhibitors, and carbapenems. This table corresponds to Figure 3D in the main text.

| **Antibiotic exposure** | **Outcome phenotype** | **Estimate** | **80% confidence interval** | | **95% confidence interval** | | **p-value** |
| --- | --- | --- | --- | --- | --- | --- | --- |
|  |  |  | **Lower bound** | **Upper bound** | **Lower bound** | **Upper bound** |  |
| **Fluoroquinolones** | S-R-S | -1.0 | -2.4 | 0.5 | -3.1 | 1.2 | 0.3799 |
|  | R-R-R | 2.2 | 0.6 | 3.8 | -0.3 | 4.6 | 0.0798 |
|  | **R-S-S** | **2.8** | **1.5** | **4.1** | **0.8** | **4.8** | **0.0051** |
|  | R-R-S | 1.4 | -0.1 | 3.1 | -1.0 | 3.9 | 0.2459 |
|  | **R-S-R** | **2.7** | **1.0** | **4.5** | **0.1** | **5.4** | **0.0387** |
|  | S-S-R | -0.1 | -2.1 | 2.0 | -3.2 | 3.1 | 0.9508 |
| **Beta-lactam/ Beta-lactamase inhibitors** | S-R-S | 1.7 | 0.5 | 3.0 | -0.1 | 3.6 | 0.0632 |
|  | R-R-R | 0.3 | -1.0 | 1.6 | -1.7 | 2.3 | 0.7779 |
|  | R-S-S | 0.2 | -0.9 | 1.3 | -1.5 | 1.9 | 0.8236 |
|  | R-R-S | 1.6 | 0.2 | 3.1 | -0.6 | 3.9 | 0.1476 |
|  | R-S-R | 0.6 | -0.9 | 2 | -1.6 | 2.8 | 0.6087 |
|  | S-S-R | -2.2 | -3.7 | -0.6 | -4.5 | 0.2 | 0.0723 |
| **Carbapenems** | S-R-S | 0.5 | -2.3 | 3.4 | -3.8 | 5 | 0.8191 |
|  | **R-R-R** | **10.4** | **7.4** | **13.6** | **5.8** | **15.3** | **< 0.0001** |
|  | R-S-S | 2.9 | -0.1 | 5.9 | -1.6 | 7.5 | 0.2107 |
|  | R-R-S | 3.1 | -0.2 | 6.6 | -2 | 8.5 | 0.2334 |
|  | **R-S-R** | **10.1** | **6.5** | **14.0** | **4.6** | **16** | **0.0003** |
|  | **S-S-R** | **15.7** | **11.5** | **20.0** | **9.4** | **22.4** | **< 0.0001** |

Table S18. Multinomial logistic regression results for *Pseudomonas aeruginosa* patchwork dataset for community prevalence covariates. Estimates are presented as the percentage change in the odds of each resistance phenotype for each increase in community prevalence of the specified antimicrobial class in the preceding 14 days, compared with isolates fully susceptible to all key classes (S-S-S phenotype). Community prevalence was measured by the incidence rate of phenotypes collected within 3 days of admission during the previous 90 days (referred to as importation phenotype). Antibiotic classes used for phenotypes: fluoroquinolones, beta-lactam/beta-lactamase inhibitors, and carbapenems. This table corresponds to Figure S3.

| **Outcome phenotype** | **Importation phenotype** | **Estimate** | **80% confidence interval** | | **95% confidence interval** | | **p-value** |
| --- | --- | --- | --- | --- | --- | --- | --- |
|  |  |  | **Lower bound** | **Upper bound** | **Lower bound** | **Upper bound** |  |
| S-R-S | S-S-S | -2.7 | -3 | -2.4 | -3.1 | -2.2 | < 0.0001 |
|  | S-R-S | 4.8 | 4.4 | 5.2 | 4.2 | 5.4 | < 0.0001 |
|  | R-R-R | 1.1 | 0.5 | 1.7 | 0.2 | 2 | 0.0153 |
|  | R-S-S | -3.2 | -3.7 | -2.6 | -4 | -2.3 | < 0.0001 |
|  | R-R-S | 1.1 | 0.6 | 1.5 | 0.3 | 1.8 | 0.0038 |
|  | R-S-R | -3.3 | -4.2 | -2.3 | -4.7 | -1.8 | < 0.0001 |
|  | S-S-R | -3 | -4.1 | -1.8 | -4.7 | -1.2 | 0.0010 |
| R-R-R | S-S-S | -1.9 | -2.2 | -1.6 | -2.4 | -1.4 | < 0.0001 |
|  | S-R-S | 3.3 | 2.8 | 3.7 | 2.6 | 4 | < 0.0001 |
|  | R-R-R | 3.4 | 2.8 | 4 | 2.5 | 4.3 | < 0.0001 |
|  | R-S-S | -1.9 | -2.5 | -1.4 | -2.7 | -1.1 | < 0.0001 |
|  | R-R-S | 2.3 | 1.8 | 2.9 | 1.6 | 3.1 | < 0.0001 |
|  | R-S-R | -1.3 | -2.2 | -0.5 | -2.6 | -0.1 | 0.0410 |
|  | S-S-R | -1 | -2.1 | 0.2 | -2.7 | 0.8 | 0.2832 |
| R-S-S | S-S-S | -0.5 | -0.8 | -0.1 | -0.9 | 0 | 0.0701 |
|  | S-R-S | -0.3 | -0.9 | 0.3 | -1.2 | 0.6 | 0.5208 |
|  | R-R-R | 0.9 | 0.1 | 1.6 | -0.2 | 2 | 0.1258 |
|  | R-S-S | 1 | 0.5 | 1.4 | 0.3 | 1.6 | 0.0034 |
|  | R-R-S | -0.2 | -0.8 | 0.5 | -1.2 | 0.9 | 0.7481 |
|  | R-S-R | 0.5 | -0.2 | 1.2 | -0.5 | 1.5 | 0.3550 |
|  | S-S-R | -0.2 | -1.1 | 0.7 | -1.6 | 1.2 | 0.7826 |
| R-R-S | S-S-S | -1.8 | -2.2 | -1.4 | -2.4 | -1.2 | < 0.0001 |
|  | S-R-S | 4.2 | 3.7 | 4.6 | 3.4 | 4.9 | < 0.0001 |
|  | R-R-R | 3.1 | 2.4 | 3.7 | 2 | 4.1 | < 0.0001 |
|  | R-S-S | -0.6 | -1.2 | -0.1 | -1.5 | 0.3 | 0.1609 |
|  | R-R-S | 4.9 | 4.4 | 5.4 | 4.1 | 5.7 | < 0.0001 |
|  | R-S-R | -1.1 | -2.1 | -0.1 | -2.6 | 0.5 | 0.1786 |
|  | S-S-R | -0.5 | -1.8 | 0.9 | -2.5 | 1.6 | 0.6377 |
| R-S-R | S-S-S | 0.1 | -0.3 | 0.5 | -0.5 | 0.8 | 0.7359 |
|  | S-R-S | -0.3 | -1.2 | 0.5 | -1.6 | 1 | 0.6155 |
|  | R-R-R | 0.1 | -0.8 | 1 | -1.3 | 1.5 | 0.9198 |
|  | R-S-S | 0 | -0.6 | 0.6 | -1 | 0.9 | 0.9625 |
|  | R-R-S | 1.2 | 0.4 | 2.1 | 0 | 2.5 | 0.0530 |
|  | R-S-R | 2.2 | 1.4 | 3 | 1 | 3.4 | 0.0005 |
|  | S-S-R | 1.3 | 0.2 | 2.5 | -0.5 | 3.2 | 0.1438 |
| S-S-R | S-S-S | -0.6 | -1 | -0.1 | -1.2 | 0.1 | 0.0932 |
|  | S-R-S | -1.6 | -2.6 | -0.5 | -3.1 | 0 | 0.0571 |
|  | R-R-R | -0.4 | -1.5 | 0.6 | -2 | 1.1 | 0.5749 |
|  | R-S-S | -0.3 | -1 | 0.3 | -1.3 | 0.7 | 0.5323 |
|  | R-R-S | -2 | -3.3 | -0.7 | -4 | 0 | 0.0520 |
|  | R-S-R | -1 | -2.1 | 0.1 | -2.6 | 0.7 | 0.2401 |
|  | S-S-R | 2.2 | 1 | 3.4 | 0.4 | 4 | 0.0151 |

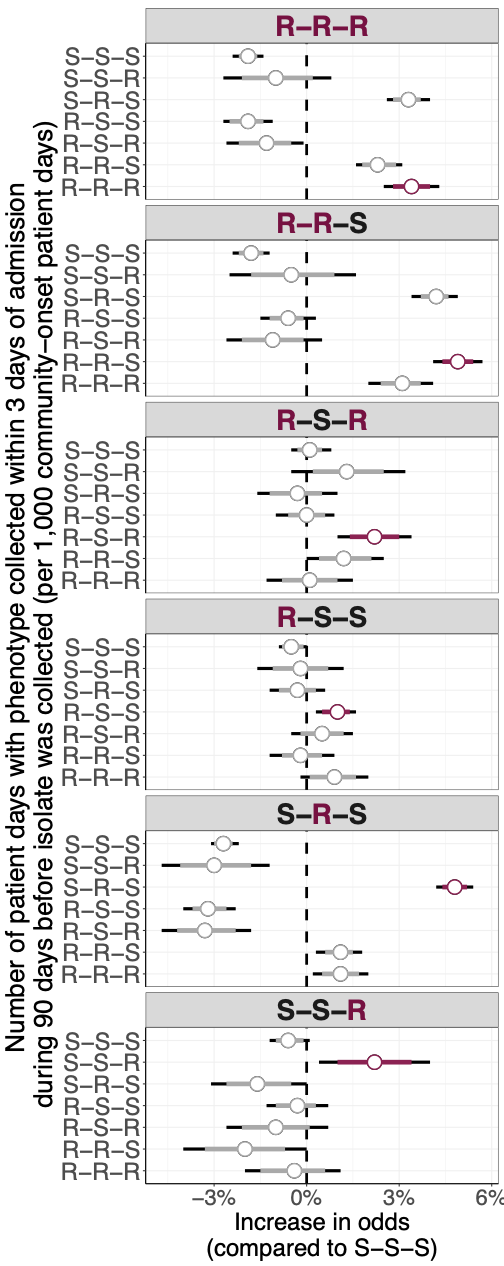

Figure S4. Multinomial logistic regression results for community prevalence covariates for *P aeruginosa*. Points represent coefficient estimates and line intervals represent confidence intervals (80% in red and 95% in black). Estimates are presented as the percentage change in the odds of each resistance phenotype for each increase in community prevalence of the specified antimicrobial class in the preceding 14 days, compared with isolates fully susceptible to all key classes (S-S-S phenotype). Community prevalence was measured by the incidence rate of phenotypes collected within 3 days of admission during the previous 90 days (referred to as importation phenotype). Antibiotic classes used for phenotypes: fluoroquinolones, beta-lactam/beta-lactamase inhibitors, and carbapenems.

#### S3.5 Adjustment for differential prescribing volumes across antimicrobial classes

In the main text, the effect of antimicrobial prescribing on the relative odds of resistance phenotypes was expressed as the percentage change in odds associated with **one additional treatment day** (per 100 patient-days) of exposure to a given antimicrobial class in the 14 days preceding isolate collection, relative to isolates fully susceptible to all key antimicrobial classes (S-S-S). However, one additional treatment day represents a different proportion of the overall prescribing rate depending on the antimicrobial class. To facilitate comparison across classes, we re-expressed the results as the percentage change in odds associated with a 1% increase in the average prescribing rate (per 100 patient-days) over the prior 14 days for each antimicrobial class. The relative effect estimates remained consistent, except for fluoroquinolones and carbapenems, whose effects became more comparable on this scale (whereas carbapenems had appeared to have a disproportionately higher effect before rescaling).

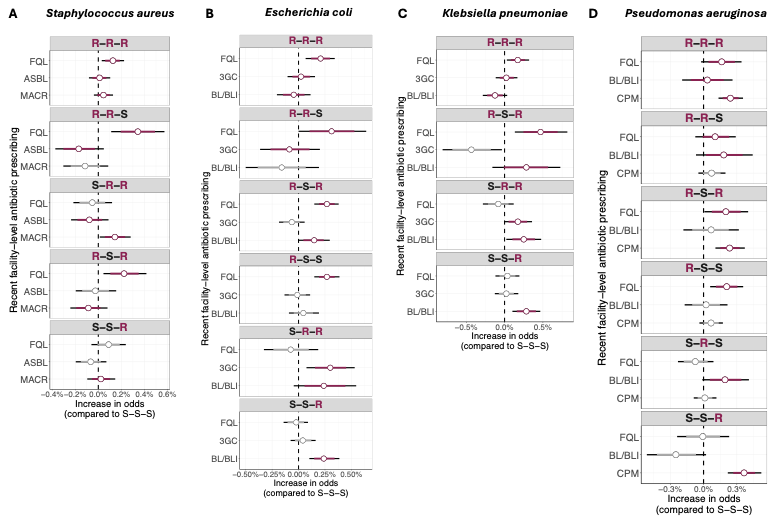

Figure S7. Rescaled effect of recent antimicrobial prescribing on resistance patterns in four bacterial pathogens. Patchwork data was used for the analysis. Multilevel multinomial logistic regression coefficients are shown as the percentage change in the odds of each resistance phenotype for a 1% increase in the average antibiotic prescribing of the specified antimicrobial class in the preceding 14 days (per 1,000 patient-days), compared with isolates fully susceptible to all key classes (S-S-S). Points denote medians; thick bars, 80 % confidence intervals; thin bars, 95 % confidence intervals. Coefficients highlighted in dark red correspond to antimicrobial-pathogen combinations for which an effect of prescribing on resistance was hypothesised. Antibiotic classes for each pathogen: (A) *S aureus:* fluoroquinolones, anti-staphylococcal beta-lactams, macrolides. (B) *E coli:* fluoroquinolones, 3^rd^ generation cephalosporins, beta-lactam/beta-lactamase inhibitors. (C) *K pneumoniae:* fluoroquinolones, 3^rd^ generation cephalosporins, beta-lactam/beta-lactamase inhibitors. (D) *P aeruginosa:* fluoroquinolones, beta-lactam/beta-lactamase inhibitors, carbapenems. FQL = fluoroquinolones, ASBL = anti-staphylococcal beta-lactams, MACR = macrolides, 3GC = 3^rd^ generation cephalosporins, BL/BLI = beta-lactam/beta-lactamase inhibitor, CPM = carbapenem

#### S3.6 Predicted probabilities of resistance phenotypes by fitted model

Predicted probabilities from the fitted model show that increasing fluoroquinolone prescribing is associated with higher probabilities of phenotypes resistant to fluoroquinolones, particularly the R–R–R, R–R–S, and R-S-R profiles. Conversely, the probability of fully susceptible (S–S–S) isolates decreases with higher FQL use, as does the probability of partially susceptible profiles (S-R-R).

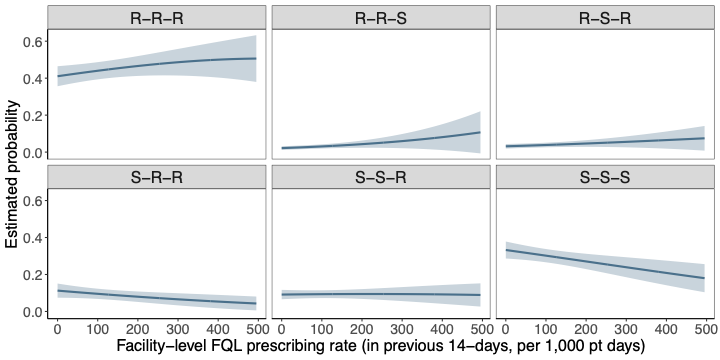

Figure S8. Predicted probabilities of *Staphylococcus aureus* resistance phenotypes as a function of recent fluoroquinolone (FQL) prescribing at the facility-level, based on the fitted multinomial logistic regression model used in the main analysis. The x-axis indicates the rate of FQL prescribing in the 14 days prior to isolate collection (per 1,000 patient-days). Each panel corresponds to a distinct resistance phenotype based on susceptibility to fluoroquinolones, antistaphylococcal beta-lactams, and macrolides. Lines represent the predicted probabilities, shaded areas the corresponding 95% confidence intervals.

#### S3.7 Single antimicrobial-single pathogen correlations

To explore unadjusted associations between facility-level antibiotic use and the prevalence of antimicrobial resistance, we examined scatterplots depicting antibiotic days of therapy in one year versus the proportion of resistant isolates collected in the next year across Veterans Affairs medical centers (Figure S5). Additionally, we summarized these associations using Spearman correlation coefficients with 95% bootstrap confidence intervals across three selected time periods (Figure S6). Together, these descriptive analyses provide a preliminary view of ecological correlations for individual pathogen–antibiotic class combinations. Most unadjusted associations were weak and varied across years, underscoring the importance of adjusting for potential confounders and temporal trends in subsequent modelling.

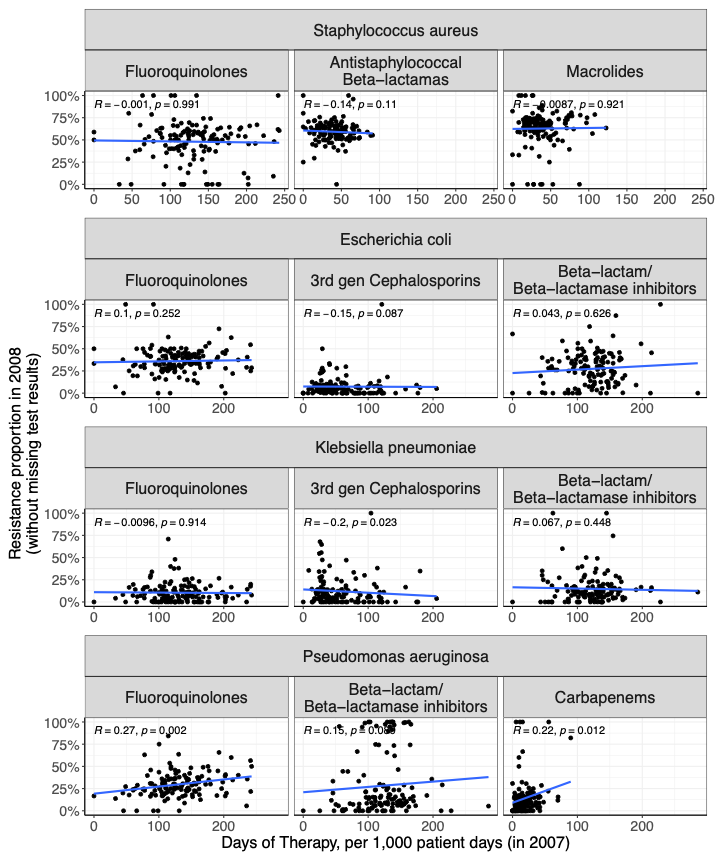

Figure S5. Scatterplots depicting single antimicrobial-pathogen use-resistance relationships. Facility-level-level antibiotic use (x-axis) was measured as days of therapy per 1,000 patient-days and measured in 2007. Resistance proportion was defined as the number of intermediate and resistant isolates divided by the total number of isolates with reported susceptibility results in 2008, among isolates collected more than 2 days after admission. Each point represents a VA facility for that year combination. The blue line represents a linear regression fit. The Spearman correlation coefficient and corresponding p-value are reported in each panel.

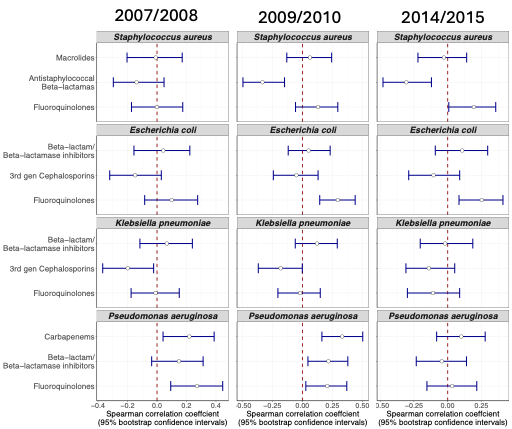

Figure S6. Unadjusted association between facility-level antibiotic use and resistance proportion for three selected time periods across single target antimicrobial-single pathogen combinations. The panels display Spearman correlation coefficients with 95% bootstrap confidence intervals, assessing the relationship between facility-level antibiotic days of therapy per 1,000 patient-days in a given year and the proportion of resistant isolates in the subsequent year. Each point indicates the estimated Spearman rank correlation coefficient, with horizontal blue lines denoting the 95% confidence intervals. Rows correspond to single pathogen–antibiotic class combinations, and columns to the three time periods (2007/2008, 2009/2010, and 2014/2015).

### S4 **Antimicrobial prescribing time trends**

We conducted time trend analyses of antibiotic prescribing in the Veterans Affairs Healthcare system by applying robust Poisson regression models with generalized estimating equations (GEE) to estimate the average annual percentage change (AAPC) in days of therapy per 1,000 patient-days for each antibiotic class. The outcome was the annual number of days of therapy, with patient-days per year included as an offset term. Models were adjusted for facility-level covariates (census region, facility rurality, facility complexity, race/ethnicity and age distribution, median length-of-stay) to account for potential confounding. We selected breakpoints based on events hypothesized to influence prescribing patterns: the onset of the COVID-19 pandemic (2020) for anti-staphylococcal beta-lactams, macrolides, 3rd-generation cephalosporins, and fluoroquinolones; a drug supply shortage in 2015 for piperacillin-tazobactam; and a VA National Antimicrobial Stewardship Initiatives in 2011 for carbapenems. Interaction terms were tested to detect significant changes in trends across these periods. Different time trend estimates were reported for respective time periods if the interaction term was statistically significant, with an overall significance level of 0.05 but corrected for multiple hypothesis testing using the Bonferroni correction method.

In 2015, the United States experienced a major shortage of piperacillin-tazobactam (commonly known by the brand name Zosyn), a widely used broad-spectrum beta-lactam/beta-lactamase inhibitor combination.^3–5^ The shortage stemmed from manufacturing disruptions, including quality control issues at facilities producing the active pharmaceutical ingredient. As a result, hospitals faced limited supplies for months, forcing clinicians to substitute other antibiotics such as cefepime, meropenem, or other beta-lactams.^4,5^

Table S19. Time trend estimates for antibiotic prescribing in the Veterans Affairs Healthcare system, Feb 1, 2007 - March 31, 2022. Estimates represent the average annual percentage change (AAPC) and were computed by performing a generalized estimating equation approach. This table corresponds to Figure 2B in the main text.

| **Antibiotic class** | **Time period** | **Time trend estimate (AAPC)** | **95% confidence interval** | | **p-value** |
| --- | --- | --- | --- | --- | --- |
|  |  |  | **Lower bound** | **Upper bound** |  |
| Anti-Staphylococcal  Beta-lactams | 2007-2019 | -1 | -1.7 | -0.2 | 0.0103 |
|  | 2020-2022 | 4.6 | 1.5 | 7.8 | 0.0038 |
| Macrolides | 2007-2019 | -0.1 | -1 | 0.9 | 0.8488 |
|  | 2020-2022 | -17.9 | -21 | -14.7 | < 0.0001 |
| Beta-lactam/Beta-lactamase inhibitors | 2007-2015 | -0.1 | -0.8 | 0.6 | 0.7993 |
|  | 2016-2022 | -2.3 | -3.9 | -0.7 | 0.0046 |
| 3^rd^ generation cephalosporins | 2007-2019 | 2.4 | 1.3 | 3.5 | < 0.0001 |
|  | 2020-2022 | -5 | -7.7 | -2.2 | 0.0005 |
| 4^th^ generation cephalosporins | 2007-2022 | 5.1 | 3.2 | 7.0 | < 0.0001 |
| 3^rd^ /4^th^ generation cephalosporins | 2007-2019 | 3.1 | 2.1 | 4.1 | < 0.0001 |
|  | 2020-2022 | -2.4 | -5.2 | 0.5 | 0.1071 |
| Carbapenems | 2007-2011 | 5.3 | 3.1 | 7.6 | < 0.0001 |
|  | 2012-2022 | -3.7 | -6.2 | -1.1 | 0.0062 |
| Fluoroquinolones | 2007-2022 | -8.4 | -9.1 | -7.7 | < 0.0001 |

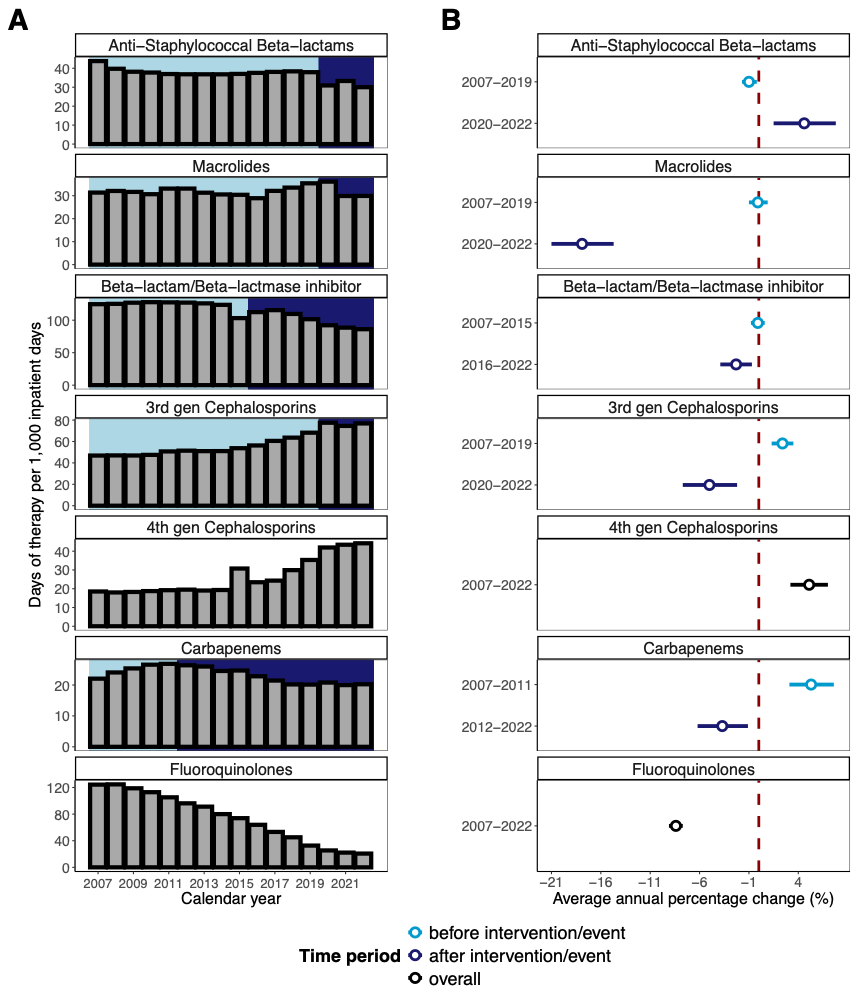

Figure S9. Antimicrobial prescribing time trends in the Veterans Affairs Healthcare Administration between 1 Feb 2007 - 31 March 2022 for key antimicrobial classes. (A) Bar plots show the overall antimicrobial prescribing rates, measured as days of therapy per 1,000 patient-days, from 1 Feb 2007 till 31 March 2022 in 138 VA medical centres for key antimicrobial classes. (B) Average annual percentage change (AAPC) estimates obtained from time trend analyses using generalized estimating equations for the antimicrobial classes shown in (A). An AAPC greater than zero indicates an increasing trend, whereas an AAPC less than zero indicates a decreasing trend. For antistaphylococcal beta-lactams, macrolides, 3rd-generation cephalosporins, and fluoroquinolones, interactions with the COVID-19 period were tested, and separate time trend estimates are displayed if the interaction term was statistically significant (overall p-value < 0.05, corrected with Bonferroni correction method). For beta-lactam/beta-lactamase inhibitors and carbapenems, interactions with time periods related to a drug supply shortage in 2015 and antimicrobial stewardship initiatives in 2011, respectively, were similarly evaluated. The terms “before intervention/event” and “after intervention/event” refer to the time periods before and after the respective breakpoints and are indicated on the y-axis.

### S5 **Simulation analyses**

To assess how variability in prescribing patterns affects coefficient estimation, we conducted a simulation study using a correctly specified multilevel multinomial logistic regression model (i.e., the same model used for fitting the data). This analysis evaluated the impact of prescribing variability on four key metrics: bias, confidence interval coverage, the envelope, and the average width of confidence intervals. The motivation behind this analysis is that when prescribing variability is low, it becomes more difficult for the model to distinguish covariate effects from the intercept, potentially leading to biased or imprecise estimates.

#### S5.1 Data Generation Process

We simulated datasets where the outcome variable represented a categorical phenotype with three antibiotic susceptibility positions, modelled similarly to the empirical data from *S aureus*. We generated datasets with 70 facilities and 150 patient isolates per facility. The predictors included:

- Facility-level antibiotic prescribing rates in the 14 days prior to culture collection
- Calendar year, month and day

The *true* regression coefficients for facility-level antibiotic prescribing were drawn from a lognormal distribution with mean $=0.0025$and sd $= 0.1$– based on estimated regression coefficients estimated from the *S aureus* patchwork data set. We assumed that the regression coefficients were the same for each antibiotic class (representing the assumption of equal selection pressure).

The intercept for each phenotype category reflects the baseline log‐odds of that category relative to the reference when all other predictors are zero. They were chosen to reflect the probabilities observed in the *S aureus* patchwork dataset.

We systematically varied the **variation in facility-level antibiotic prescribing**:

We generated scenarios where prescribing rates had low, medium, or high variability across facilities to examine how the range of antibiotic use affects inference. The distributions were based on a log-normal distribution with the same mean of 117.21 (based on fluoroquinolone prescribing rates in the *S aureus* patchwork dataset) and the following parameters:

1. Low: meanlog = 4.70, sdlog = 0.36 (Drug 1)
2. Medium: meanlog = 4.50, sdlog = 0.72 (Drug 2)
3. High variability: meanlog = 3.72, sdlog = 1.44 (Drug 3)

Each simulation scenario was repeated 100 times to ensure robust results.

#### S5.2 Model Fitting and Evaluation Metrics

For each simulated dataset, we fit a **correctly specified multinomial logistic regression model** using the same structure as the empirical analysis. We evaluated:

- **Bias**: The difference between the estimated and true regression coefficients.
- **Confidence Interval Coverage**: The proportion of simulations where the true coefficient was contained within the 95% confidence interval.
- **Envelope of Confidence Intervals**: The range of observed confidence intervals across simulations, capturing the variability in uncertainty.
- **Average Confidence Interval Width**: The mean width of confidence intervals across simulations, reflecting the overall level of uncertainty in effect estimates.

### S6 **Sensitivity analyses**

In this section, we report the results of several sensitivity analyses to test the robustness of our main conclusions.

#### S6.1 Dealing with missing antimicrobial susceptibility test results

For some antibiotic classes, a substantial number of isolates have non-reported antimicrobial susceptibility test (AST) results. Since the reasons for these missing AST results are not known to us, simply excluding these isolates not only drastically reduced the sample size but also may introduce bias in the results (especially if the missing results are due to selective reporting to reduce inappropriate and unnecessary antibiotic prescribing). To overcome this challenge, we used two main approaches for each antibiotic-pathogen combination:

- Patchwork approach: For each 1-year segment, we calculated the proportion of missing AST results for each facility and included only facilities that do not exceed the threshold (i.e., 30%) in consecutive quarters. For this approach, a facility does not need to be present in all segments but only in those where it meets the criteria. We merged the subsets from each 1-year period to form a comprehensive dataset. The resulting dataset will be a composite of different facilities in different time segments. This approach will allow more facilities to be included resulting in a larger sample size.
- Consistent quality approach: We created a dataset with facilities and their isolates only included if the proportion of missing test results $\leq$ 30% for each consecutive quarter for each year. This approach results in a dataset that encompasses facilities that maintain a certain level of data completeness throughout the entire study period. However, it resulted in a very small subset of the original dataset and the included facilities may be different from the rest of the dataset.

We used the patchwork approach as our main approach for the analysis and compared the results to the results of the *consistent quality approach* and to the results of the approach where the full dataset was used with isolates with missing AST results removed. In addition, we compared these results to a third approach:

- Complete dataset with missing susceptibility test results removed: We removed missing susceptibility test results for the key antibiotic classes. For this approach, it is assumed that these missing test results are missing at random.

We performed our use-resistance analyses using the whole dataset with missing susceptibility test results removed instead of using the patchwork dataset. This approach implicitly assumes that the missing test results are missing at random.

##### S6.1.1 *Staphylococcus aureus*

Results using the complete dataset, in which isolates with missing susceptibility test results were excluded, were generally consistent with our main findings from the patchwork dataset. The primary conclusions did not materially change. Some estimates appeared more precise and further from the null, with narrower 95% confidence intervals, likely reflecting the increased sample size in the complete dataset when compared to the patchwork dataset. Notable differences were observed for phenotypes susceptible to fluoroquinolones and resistant to anti-staphylococcal beta-lactams and macrolides (S-R-R), as well as phenotypes resistant only to macrolides (S-S-R). For these phenotypes, recent prescribing of anti-staphylococcal beta-lactams and macrolides was associated with increased relative odds, respectively, although the 95% confidence intervals included zero, indicating statistical uncertainty around these estimates.

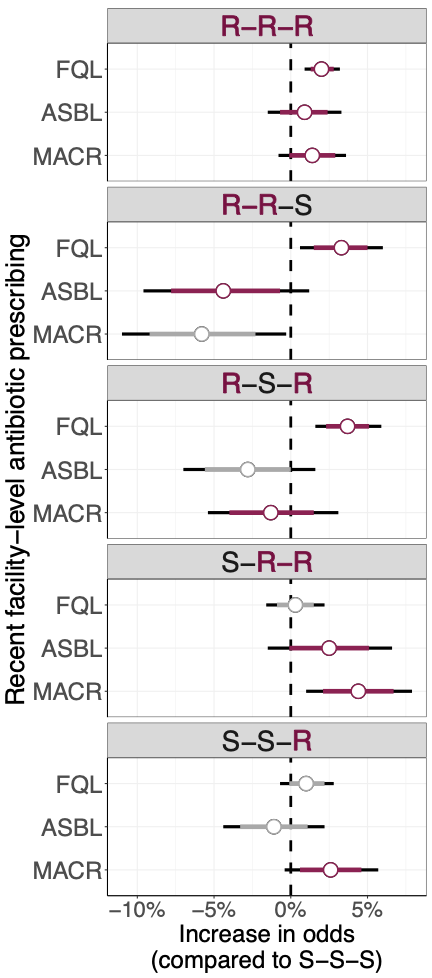

Figure S10. Effect of recent antimicrobial prescribing on resistance patterns in *Staphylococcus aureus* using the complete dataset with missing susceptibility test results removed. Multilevel multinomial logistic regression coefficients are shown as the percentage change in the odds of each resistance phenotype for every additional treatment day (per 100 patient-days) of exposure to the specified antimicrobial class in the preceding 14 days (per 1,000 patient-days), compared with isolates fully susceptible to all key classes (S-S-S). Points denote medians; thick bars, 80 % confidence intervals; thin bars, 95 % confidence intervals. Coefficients highlighted in dark red correspond to antimicrobial-pathogen combinations for which an effect of prescribing on resistance was hypothesised. Antibiotic classes for each pathogen: FQL = fluoroquinolones, ASBL = anti-staphylococcal beta-lactams, MACR = macrolides.

##### S6.1.2 *Escherichia coli*

Results using the complete dataset, in which isolates with missing susceptibility test results were excluded, were generally consistent with our main findings from the patchwork dataset. The primary conclusions did not materially change. Some estimates appeared more precise and further from the null, with narrower 95% confidence intervals, likely reflecting the increased sample size in the complete dataset when compared to the patchwork dataset.

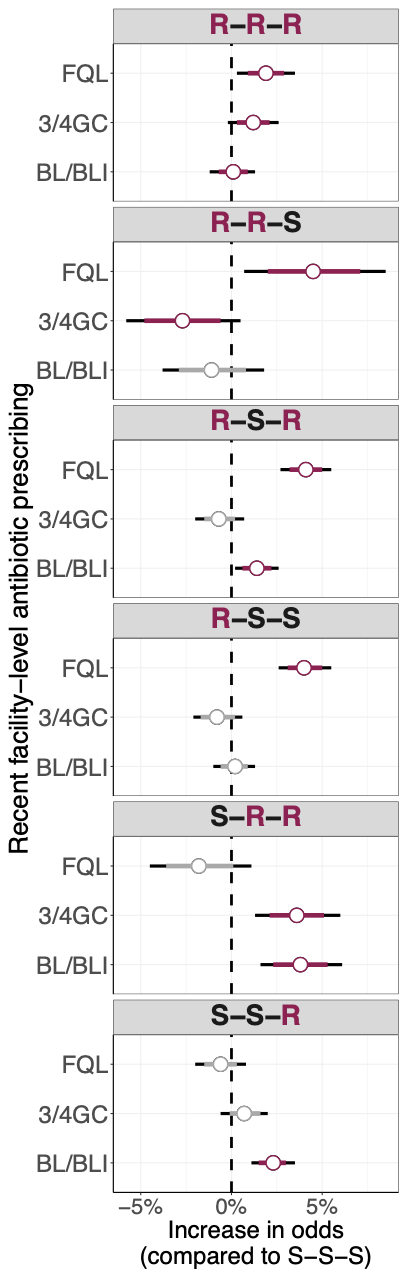

Figure S11. Effect of recent antimicrobial prescribing on resistance patterns in *Escherichia coli* using the complete dataset with missing susceptibility test results removed. Multilevel multinomial logistic regression coefficients are shown as the percentage change in the odds of each resistance phenotype for every additional treatment day (per 100 patient-days) of exposure to the specified antimicrobial class in the preceding 14 days (per 1,000 patient-days), compared with isolates fully susceptible to all key classes (S-S-S). Points denote medians; thick bars, 80 % confidence intervals; thin bars, 95 % confidence intervals. Coefficients highlighted in dark red correspond to antimicrobial-pathogen combinations for which an effect of prescribing on resistance was hypothesised. Antibiotic classes for each pathogen: FQL = fluoroquinolones, 3GC = 3^rd^ generation cephalosporins, BL/BLI = beta-lactam/beta-lactamase inhibitors.

##### S6.1.3 *Klebsiella pneumoniae*

Results using the complete dataset, in which isolates with *K pneumoniae* missing susceptibility test results for FQLs, 3/4GCs, or BL/BLIs were excluded, were generally consistent with our main findings from the patchwork dataset. The primary conclusions did not materially change. Compared to results for other pathogens, estimates from the complete dataset were not necessarily more precise than those from the patchwork data. For the S-R-R phenotype, 3/4GC prescribing showed a smaller median estimate. The larger sample size in the complete dataset allowed us to additionally examine the R-S-S phenotype, which revealed that fluoroquinolone prescribing increased the relative odds of the FQL-resistant, 3GC- and BL/BLI-susceptible *Klebsiella pneumoniae* phenotype.

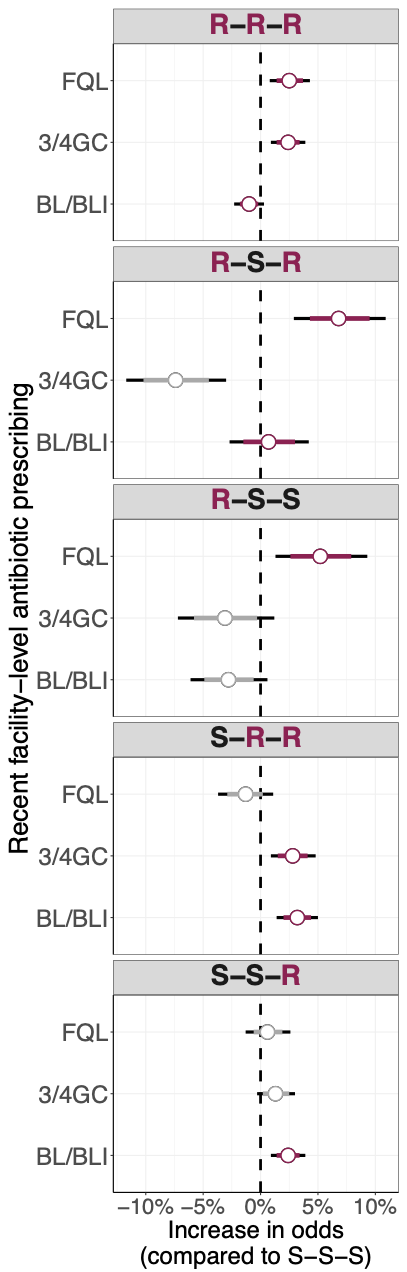

Figure S12. Effect of recent antimicrobial prescribing on resistance patterns in *Klebsiella pneumoniae* using the complete dataset with missing susceptibility test results removed. Multilevel multinomial logistic regression coefficients are shown as the percentage change in the odds of each resistance phenotype for every additional treatment day (per 100 patient-days) of exposure to the specified antimicrobial class in the preceding 14 days (per 1,000 patient-days), compared with isolates fully susceptible to all key classes (S-S-S). Points denote medians; thick bars, 80 % confidence intervals; thin bars, 95 % confidence intervals. Coefficients highlighted in dark red correspond to antimicrobial-pathogen combinations for which an effect of prescribing on resistance was hypothesised. Antibiotic classes for each pathogen: FQL = fluoroquinolones, 3GC = 3^rd^ generation cephalosporins, BL/BLI = beta-lactam/beta-lactamase inhibitors.

##### S6.1.4 *Pseudomonas aeruginosa*

Results using the complete dataset, in which *P aeruginosa* isolates with missing susceptibility test results for FQL, BL/BLIs, or CPMs were excluded, were generally consistent with our main findings from the patchwork dataset. The primary conclusions did not materially change (Figure S11). For the R-R-S phenotype, the estimates for the effect of antibiotic prescribing on resistance patterns were more precise and did not include the null. For other phenotypes and their corresponding antibiotic classes, the point estimates were generally smaller compared to those obtained from the patchwork dataset.

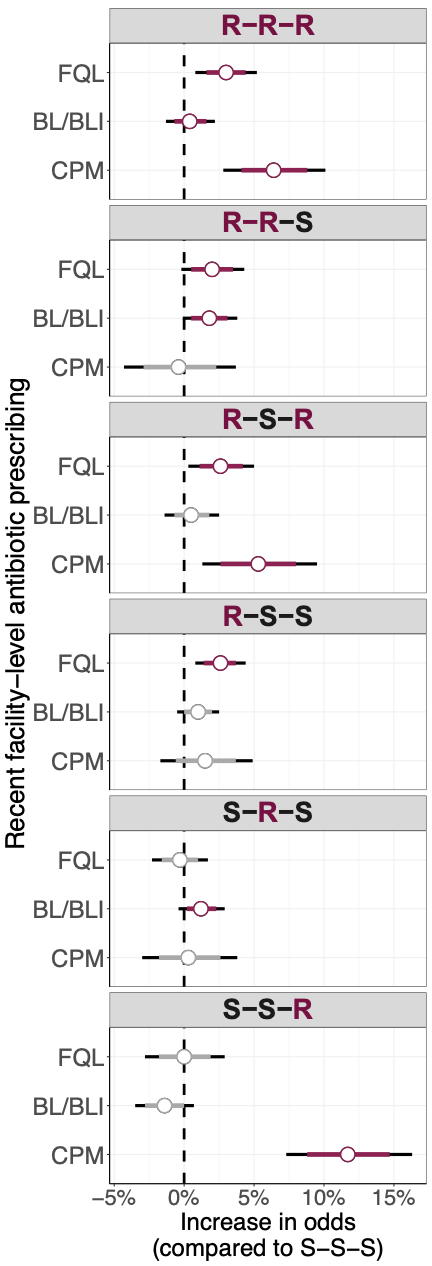

Figure S13. Effect of recent antimicrobial prescribing on resistance patterns in *Pseudomonas aeruginosa* using the complete dataset with missing susceptibility test results removed. Multilevel multinomial logistic regression coefficients are shown as the percentage change in the odds of each resistance phenotype for every additional treatment day (per 100 patient-days) of exposure to the specified antimicrobial class in the preceding 14 days (per 1,000 patient-days), compared with isolates fully susceptible to all key classes (S-S-S). Points denote medians; thick bars, 80 % confidence intervals; thin bars, 95 % confidence intervals. Coefficients highlighted in dark red correspond to antimicrobial-pathogen combinations for which an effect of prescribing on resistance was hypothesised. Antibiotic classes for each pathogen: FQL = fluoroquinolones, BL/BLI = beta-lactam/beta-lactamase inhibitors, CPM = carbapenems.

#### S6.2 Extending antibiotic exposure period

In our main analysis, we defined antibiotic exposure as prescriptions issued within 14 days prior to the date the phenotype was isolated. To assess the robustness of this choice, we conducted a sensitivity analysis extending the exposure window to 30 days before the isolation date and evaluated whether the results were materially affected. The results and overall conclusions remained consistent, although the estimates generally showed greater uncertainty (Figure S 12).

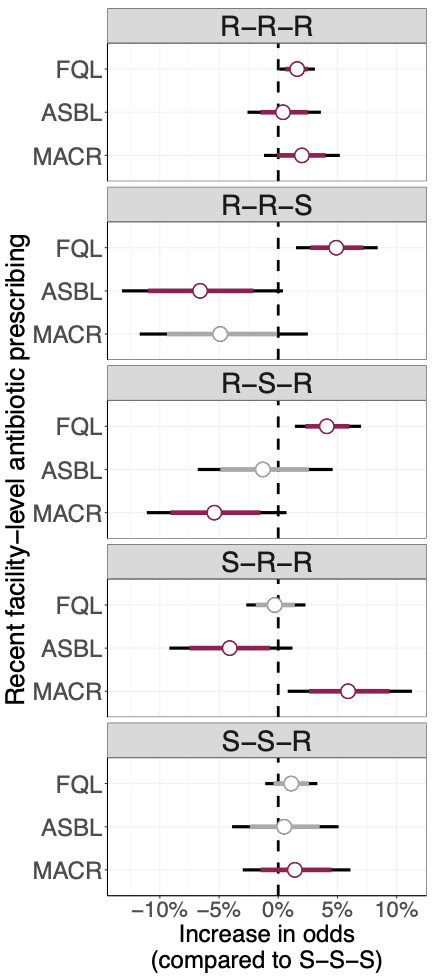

Figure S14. Effect of recent antimicrobial prescribing on resistance patterns in *Staphylococcus aureus* using a 30-day exposure window. Patchwork data with an extended antibiotic exposure window (30 days) were used for this analysis. Multilevel multinomial logistic regression coefficients are shown as the percentage change in the odds of each resistance phenotype for every additional treatment day (per 100 patient-days) of exposure to the specified antimicrobial class in the preceding 14 days (per 1,000 patient-days), compared with isolates fully susceptible to all key classes (S-S-S). Points denote medians; thick bars, 80 % confidence intervals; thin bars, 95 % confidence intervals. Coefficients highlighted in dark red correspond to antimicrobial-pathogen combinations for which an effect of prescribing on resistance was hypothesised. Antibiotic classes for each pathogen: FQL = fluoroquinolones, ASBL = anti-staphylococcal beta-lactams, MACR = macrolides.

#### S6.3 Accounting for Past Incidence of Hospital-onset Phenotypes

We included an additional covariate representing the number of hospital-onset phenotypes in the 14 days prior to facility-level antibiotic treatment (per 1,000 patient days). Results did not change considerably when compared to results from our main analysis (Figure S 13).

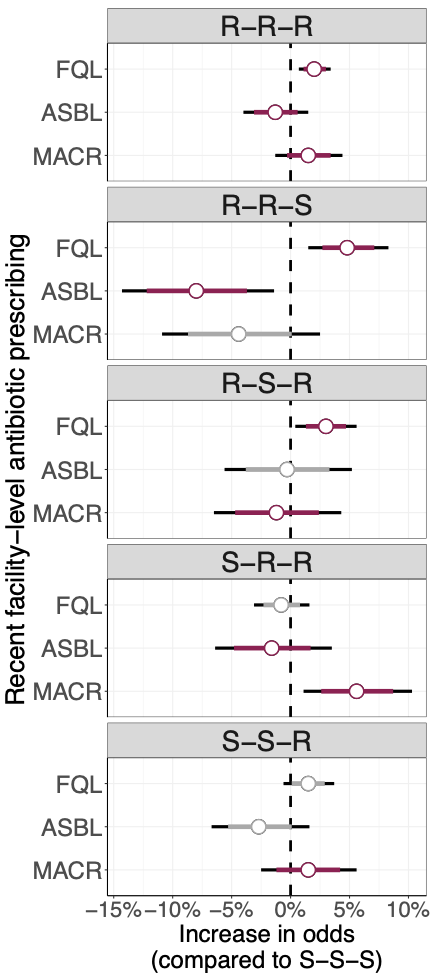

Figure S15. Effect of recent antimicrobial prescribing on resistance patterns in *Staphylococcus aureus* with additional covariates representing past facility-level incidence of hospital-onset phenotypes. Patchwork data was used for this analysis. Multilevel multinomial logistic regression coefficients are shown as the percentage change in the odds of each resistance phenotype for every additional treatment day (per 100 patient-days) of exposure to the specified antimicrobial class in the preceding 14 days (per 1,000 patient-days), compared with isolates fully susceptible to all key classes (S-S-S). Points denote medians; thick bars, 80 % confidence intervals; thin bars, 95 % confidence intervals. Coefficients highlighted in dark red correspond to antimicrobial-pathogen combinations for which an effect of prescribing on resistance was hypothesised. Antibiotic classes for each pathogen: FQL = fluoroquinolones, ASBL = anti-staphylococcal beta-lactams, MACR = macrolides.

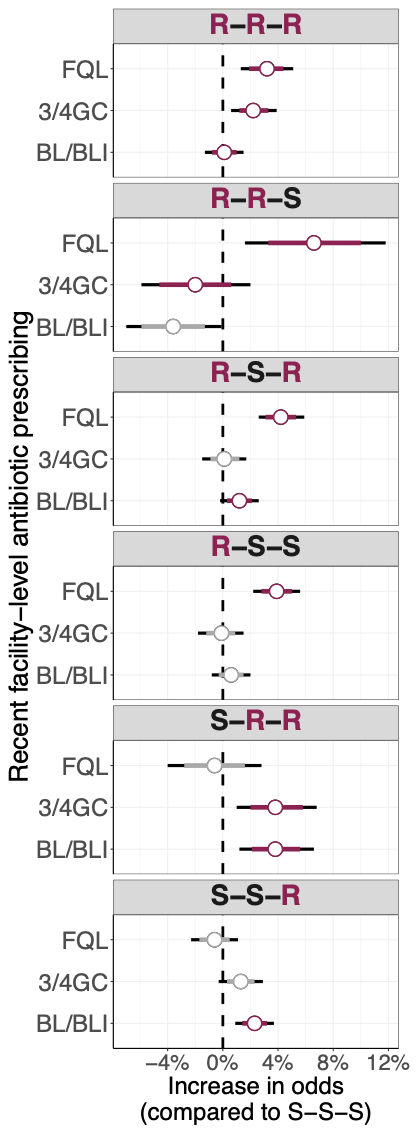

Figure S 16. Effect of recent antimicrobial prescribing on resistance patterns in *Escherichia coli* with additional covariates representing past facility-level incidence of hospital-onset phenotypes. Patchwork data was used for this analysis. Multilevel multinomial logistic regression coefficients are shown as the percentage change in the odds of each resistance phenotype for every additional treatment day (per 100 patient-days) of exposure to the specified antimicrobial class in the preceding 14 days (per 1,000 patient-days), compared with isolates fully susceptible to all key classes (S-S-S). Points denote medians; thick bars, 80 % confidence intervals; thin bars, 95 % confidence intervals. Coefficients highlighted in dark red correspond to antimicrobial-pathogen combinations for which an effect of prescribing on resistance was hypothesised. Antibiotic classes for each pathogen: FQL = fluoroquinolones, 3/4GC = 3^rd^ and 4^th^ generation cephalosporins, BL/BLI = Beta-lactam/Beta-lactamase inhibitors.

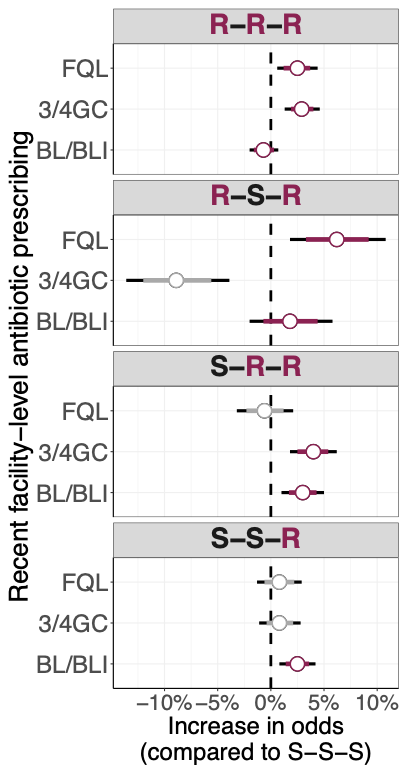

Figure S17. Effect of recent antimicrobial prescribing on resistance patterns in *Klebsiella pneumoniae* with additional covariates representing past facility-level incidence of hospital-onset phenotypes. Patchwork data was used for this analysis. Multilevel multinomial logistic regression coefficients are shown as the percentage change in the odds of each resistance phenotype for every additional treatment day (per 100 patient-days) of exposure to the specified antimicrobial class in the preceding 14 days (per 1,000 patient-days), compared with isolates fully susceptible to all key classes (S-S-S). Points denote medians; thick bars, 80 % confidence intervals; thin bars, 95 % confidence intervals. Coefficients highlighted in dark red correspond to antimicrobial-pathogen combinations for which an effect of prescribing on resistance was hypothesised. Antibiotic classes for each pathogen: FQL = fluoroquinolones, 3/4GC = 3^rd^ and 4^th^ generation cephalosporins, BL/BLI = Beta-lactam/Beta-lactamase inhibitors.

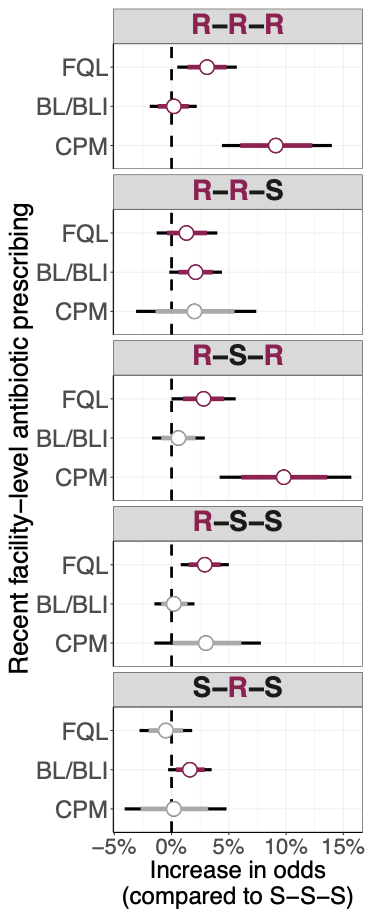

Figure S18. Effect of recent antimicrobial prescribing on resistance patterns in *Pseudomonas aeruginosa* with additional covariates representing past facility-level incidence of hospital-onset phenotypes. Patchwork data was used for this analysis. Multilevel multinomial logistic regression coefficients are shown as the percentage change in the odds of each resistance phenotype for every additional treatment day (per 100 patient-days) of exposure to the specified antimicrobial class in the preceding 14 days (per 1,000 patient-days), compared with isolates fully susceptible to all key classes (S-S-S). Points denote medians; thick bars, 80 % confidence intervals; thin bars, 95 % confidence intervals. Coefficients highlighted in dark red correspond to antimicrobial-pathogen combinations for which an effect of prescribing on resistance was hypothesised. Antibiotic classes for each pathogen: FQL = fluoroquinolones, BL/BLI = Beta-lactam/Beta-lactamase inhibitors, CPM = antipseudomonal carbapenems.

#### S6.4 Inclusion of Vancomycin Exposure for *Staphylococcus aureus*

For *S aureus,* we included an additional covariate capturing facility-level prescribing of vancomycin. This adjustment aimed to account for potential reverse confounding: facilities with a high prevalence of MRSA might avoid prescribing anti-staphylococcal beta-lactams (ASBL), which could bias the observed association toward a protective effect of ASBL. Because facilities with high MRSA prevalence are more likely to prescribe vancomycin, including vancomycin exposure in the model could help adjust for this practice pattern. However, the inclusion of this covariate did not materially change the results or main conclusions (Figure S14). In addition, we did not observe an effect of vancomycin prescribing on the included resistance phenotypes.

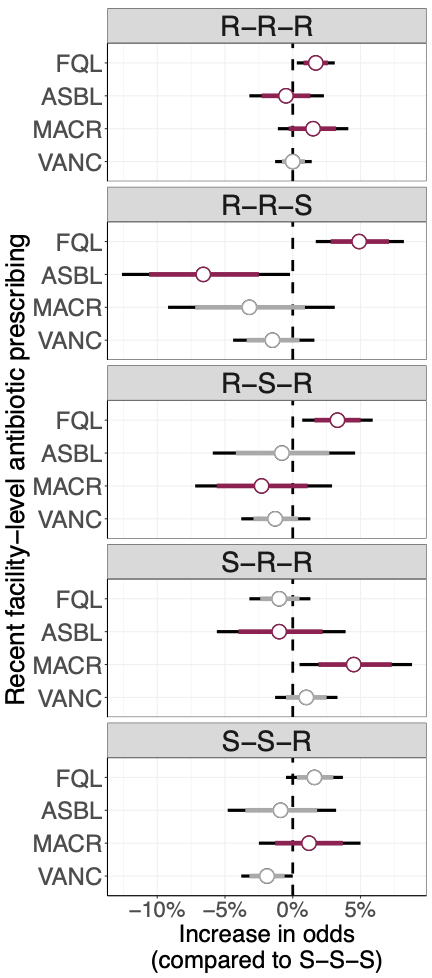

Figure S19. Effect of recent antimicrobial prescribing on resistance patterns in *Staphylococcus aureus* with additional covariate representing facility-level vancomycin prescribing in the past 14 days. Patchwork data was used for this analysis. Multilevel multinomial logistic regression coefficients are shown as the percentage change in the odds of each resistance phenotype for every additional treatment day (per 100 patient-days) of exposure to the specified antimicrobial class in the preceding 14 days (per 1,000 patient-days), compared with isolates fully susceptible to all key classes (S-S-S). Points denote medians; thick bars, 80 % confidence intervals; thin bars, 95 % confidence intervals. Coefficients highlighted in dark red correspond to antimicrobial-pathogen combinations for which an effect of prescribing on resistance was hypothesised. Antibiotic classes for each pathogen: FQL = fluoroquinolones, ASBL = anti-staphylococcal beta-lactams, MACR = macrolides.

#### S6.5 Interaction between antibiotic use variables

For *E coli and K pneumoniae,* an interaction term between 3GC and FQL use was tested but excluded from the final model due to lack of statistical significance and model fit improvement.
